# Using multivariable Mendelian randomization to estimate the causal effect of bone mineral density on osteoarthritis risk, independent of body mass index

**DOI:** 10.1101/2021.03.22.21253803

**Authors:** April Hartley, Eleanor Sanderson, Raquel Granell, Lavinia Paternoster, Jie Zheng, George Davey Smith, Lorraine Southam, Konstantinos Hatzikotoulas, Cindy G Boer, Joyce van Meurs, Eleftheria Zeggini, The Genetics of Osteoarthritis consortium, Celia L Gregson, Jon H Tobias

## Abstract

**Objectives:** Observational analyses suggest that high Bone Mineral Density (BMD) is a risk factor for osteoarthritis (OA); it’s unclear whether this represents a causal effect or shared aetiology and whether these relationships are body mass index (BMI)-independent. We performed bidirectional Mendelian randomization (MR) to uncover the causal pathways between BMD, BMI and OA.

**Methods:** One-sample (1S)MR estimates were generated by two-stage least-squares regression. Unweighted allele scores instrumented each exposure. Two-sample (2S)MR estimates were generated using inverse-variance weighted fixed-effects meta-analysis. Multivariable MR (MVMR), including BMD and BMI instruments in the same model, determined the BMI-independent causal pathway from BMD to OA. Latent causal variable (LCV) analysis, using weight-adjusted FN-BMD and hip/knee OA summary statistics, determined if genetic correlation explained the causal effect of BMD on OA.

**Results:** 1SMR provided strong evidence for a causal effect of eBMD on hip and knee OA (OR_hip_ =1.28[1.05,1.57],p=0.02, OR_knee_ =1.40[1.20,1.63],p=3×10^−5^, OR per SD increase). 2SMR effect sizes were consistent in direction. Results suggested that the causal pathways between eBMD and OA were bidirectional (β_hip_=1.10[0.36,1.84],p=0.003, β _knee_ =4.16[2.74,5.57],p=8×10^−9^, β=SD increase per doubling in risk). MVMR identified a BMI-independent causal pathway between eBMD and hip/knee OA. LCV suggested that genetic correlation (i.e. shared genetic aetiology) did not fully explain causal effects of BMD on hip/knee OA.

**Conclusions:** These results provide evidence for a BMI-independent causal effect of eBMD on OA. Despite evidence of bidirectional effects, the effect of BMD on OA did not appear to be fully explained by shared genetic aetiology, suggesting a direct action of bone on joint deterioration.

## Introduction

Although osteoarthritis (OA) is a major cause of morbidity worldwide, effective pharmacological treatment remains elusive. It may be possible to develop novel therapeutic approaches based on understanding of risk factors. Several large population-based studies have identified positive relationships between bone mineral density (BMD) and hip and knee OA (reviewed in (1)). Mendelian Randomisation (MR), which is commonly used to explore causal relationships (2, 3), has recently obtained evidence for a causal role of BMD on hip and knee OA risk (4). (BMI), a risk factor for OA (5-7) and positively associated with BMD (8), may bias MR estimates for the relationship between BMD and OA. Funck-Brentano *et al* addressed this by excluding instrument(s) associated with BMI (4). An alternative approach, yet to be applied in this context, is the use of multivariable MR (MVMR) to estimate the direct causal effect of the exposure on the outcome when the instrument(s) are associated with multiple risk factors (9). Alternatively, rather than a causal effect of BMD on OA, shared biological pathways may contribute to both traits. Consistent with this possibility, a genetic correlation between lumbar spine (LS)-BMD and OA has been observed (10). Genetic correlation may give rise to bidirectional causal relationships in MR analysis.

As well as the relationship between BMD and OA, relationships with BMI could be characterised by bidirectional relationships. A causal effect of BMI on BMD is well-established; the skeleton adapts to the increased load placed upon it, by increasing BMD. Alternatively, a causal pathway between BMD and BMI is plausible, via the metabolic effects of bone turnover. Murine osteocalcin knockouts have increased fat mass and are insulin resistant (11); in humans higher BMD is associated with lower circulating osteocalcin, which may mediate the positive association between BMD and fat mass. However, an MR analysis found no evidence of a causal pathway between femoral neck (FN) or LS-BMD and BMI in children (8).

To provide a more complete understanding of the relationship between BMD and OA, we tested bidirectional relationships between BMD, OA and BMI (*Figure 1*) using one-sample (1S) and two-sample (2S) MR, and aimed to determine the direct (*i*.*e*. unconfounded) causal pathways between these variables using MVMR.

**Figure 1:**
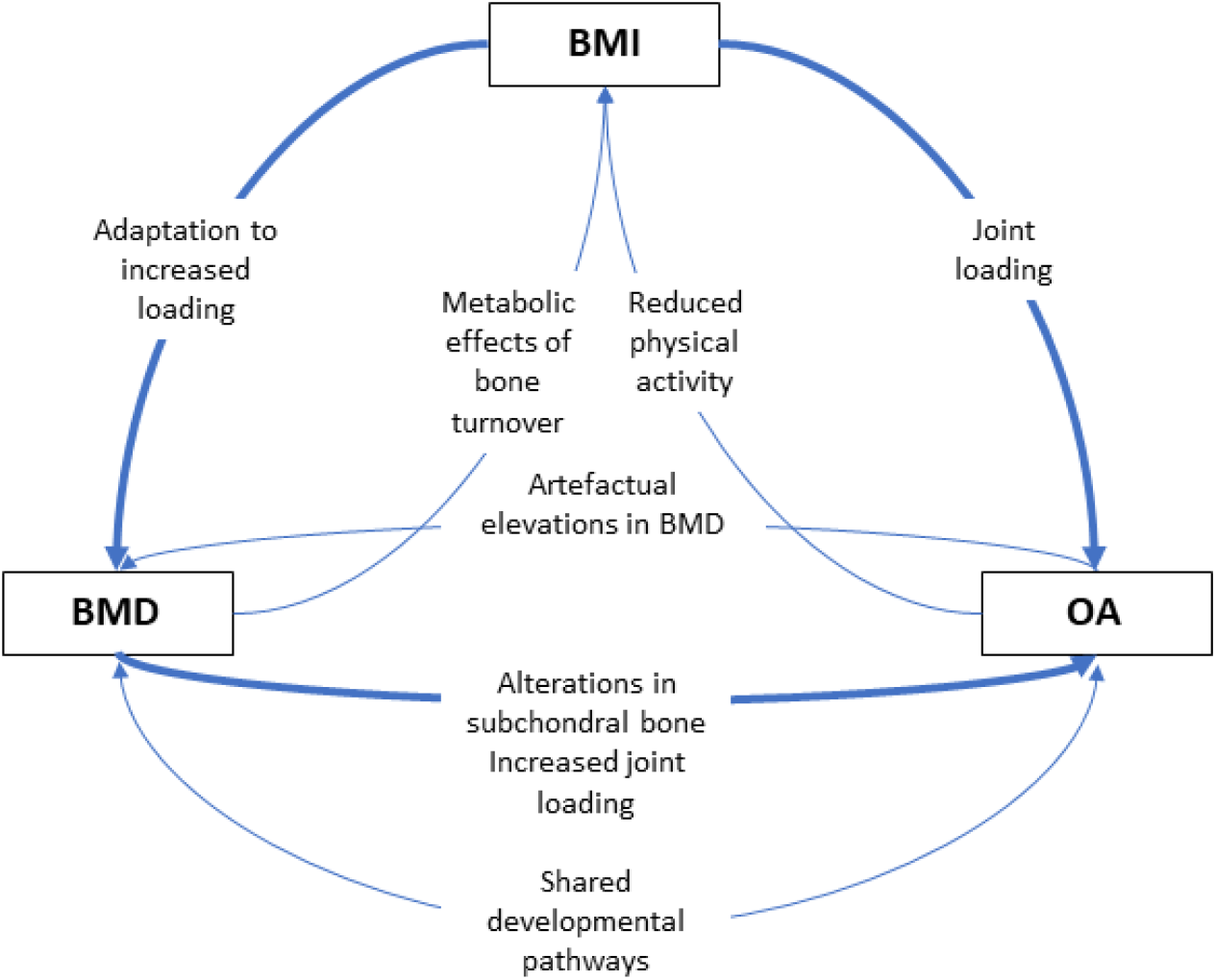
hypothesized causal diagram for relationships between BMD, BMI and OA. Thicker arrows represent stronger hypothesized relationships Abbreviations: BMD: bone mineral density; BMI: body mass index, OA: osteoarthritis

## Methods

### Individual-level analyses

Individual-level analyses were performed in the UK Biobank (UKBB) population. Data collection, genotyping and imputation and observational analyses in UK Biobank are described in the *Supplementary Material.*

### Mendelian Randomization

A summary of all MR analyses performed, along with the source of each of the instruments, is presented in Table 1 and of the assumptions of MR and how we tested these in Figure 2.

**Table 1:** summary of all one-sample and two-sample MR analyses performed

| Exposure |  |  | Outcome |  |
| --- | --- | --- | --- | --- |
|  |  | Source |  | Source |
| 1S | eBMD | Individual-level eBMD in UKBB<br>I: FN-BMD SNPs from GEFOS (19) | Knee OA | Individual-level HD knee OA status in UKBB |
| 2S | eBMD | Summary statistics from GEFOS UKBB eBMD GWAS<br>N=426 824 (28) | Knee OA | Summary statistics from GO consortium GWAS based on radiographic, clinical evaluation, joint replacement, self-reported or HD knee OA, excluding UKBB<br>N=44 001 ca, 301 541 co (29) |
| 1S | eBMD | Individual-level eBMD in UKBB<br>I: FN-BMD SNPs from GEFOS (19) | Hip OA | Individual-level HD hip OA status in UKBB |
| 2S | eBMD | Summary statistics from GEFOS UKBB eBMD GWAS<br>N=426 824 (28) | Hip OA | Summary statistics from GO consortium GWAS based on radiographic, clinical evaluation, joint replacement, self-reported or HD hip OA, excluding UKBB<br>N=25 237 ca, 272 284 co (29) |
| 1S | eBMD | Individual-level eBMD in UKBB<br>I: FN-BMD SNPs from GEFOS (19) | BMI | Individual-level BMI data in UKBB |
| 2S | eBMD | Summary statistics from GEFOS UKBB eBMD GWAS<br>N=426 824 (28) | BMI | Summary statistics from GIANT European BMI GWAS<br>N=339 224 (30) |
| 1S | BMI | Individual-level BMI data in UKBB<br>I: BMI SNPs from GIANT (30) | Knee OA | Individual-level HD knee OA status in UKBB |
| 2S | BMI | Summary statistics from GIANT European BMI GWAS<br>N=339 224 (30) | Knee OA | Summary statistics from UKBB and arcOGEN GWAS HD knee OA<br>N=24 955 ca, 378 169 co (31) |
| 1S | BMI | Individual-level BMI data in UKBB<br>I: BMI SNPs from GIANT (30) | Hip OA | Individual-level HD hip OA status in UKBB |
| 2S | BMI | Summary statistics from GIANT European BMI GWAS<br>N=339 224 (30) | Hip OA | Summary statistics from UKBB and arcOGEN GWAS of HD hip OA<br>N=15 704 ca, 378 169 co (31) |
| 1S | BMI | Individual-level BMI data in UKBB<br>I: BMI SNPs from GIANT (30) | eBMD | Individual-level eBMD data in UKBB |
| 2S | BMI | Summary statistics from GIANT European BMI GWAS<br>N=339 224 (30) | eBMD | Summary statistics from GEFOS UKBB eBMD GWAS<br>N=426 824 (28) |
| 1S | Knee OA | Individual-level data on HD knee OA in UKBB<br>I: knee OA SNPs from the GO consortium meta-analysis (excluding UKBB) (29) | eBMD | Individual-level eBMD data in UKBB |
| 2S | Knee OA | Summary statistics from GO consortium GWAS of knee OA, excluding UKBB<br>N=44 001 ca, 301 541 co (29) | eBMD | Summary statistics from GEFOS UKBB eBMD GWAS<br>N=426 824 (28) |
| 1S | Knee OA | Individual-level data on HD knee OA in UKBB<br>I: knee OA SNPs from the GO consortium meta-analysis (excluding UKBB) (29) | BMI | Individual-level BMI data in UKBB |
| 2S | Knee OA | Summary statistics from UKBB and arcOGEN GWAS of HD knee OA<br>N=24 955 ca, 378 169 co (31) | BMI | Summary statistics from GIANT European BMI GWAS<br>N=339 224 (30) |
| 1S | Hip OA | Individual-level data on HD hip OA in UKBB<br>I: hip OA SNPs from the GO consortium meta-analysis (excluding UKBB) (29) | eBMD | Individual-level eBMD data in UKBB |
| 2S | Hip OA | Summary statistics from GO consortium GWAS of hip OA, excluding UKBB<br>N=25 237 ca, 272 284 co (29) | eBMD | Summary statistics from GEFOS UKBB eBMD GWAS<br>N=426 824 (28) |
| 1S | Hip OA | Individual-level data on HD hip OA in UKBB<br>I: hip OA SNPs from the GO consortium meta-analysis (excluding UKBB) (29) | BMI | Individual-level BMI data in UKBB |
| 2S | Hip OA | Summary statistics from UKBB and arcOGEN GWAS of HD hip OA<br>N=15 704 ca, 378 169 co (31) | BMI | Summary statistics from GIANT European BMI GWAS<br>N=339 224 (30) |

*Abbreviations: 1S: one-sample MR; 2S: two-sample MR; I: instrumented by; ca: cases; co: controls; eBMD: estimated bone mineral density; BMI: Body Mass Index; HD: hospital-diagnosed; FN-BMD: femoral neck bone mineral density; OA: osteoarthritis; PCs: principal components; UKBB: UK Biobank*

**Figure 2:**
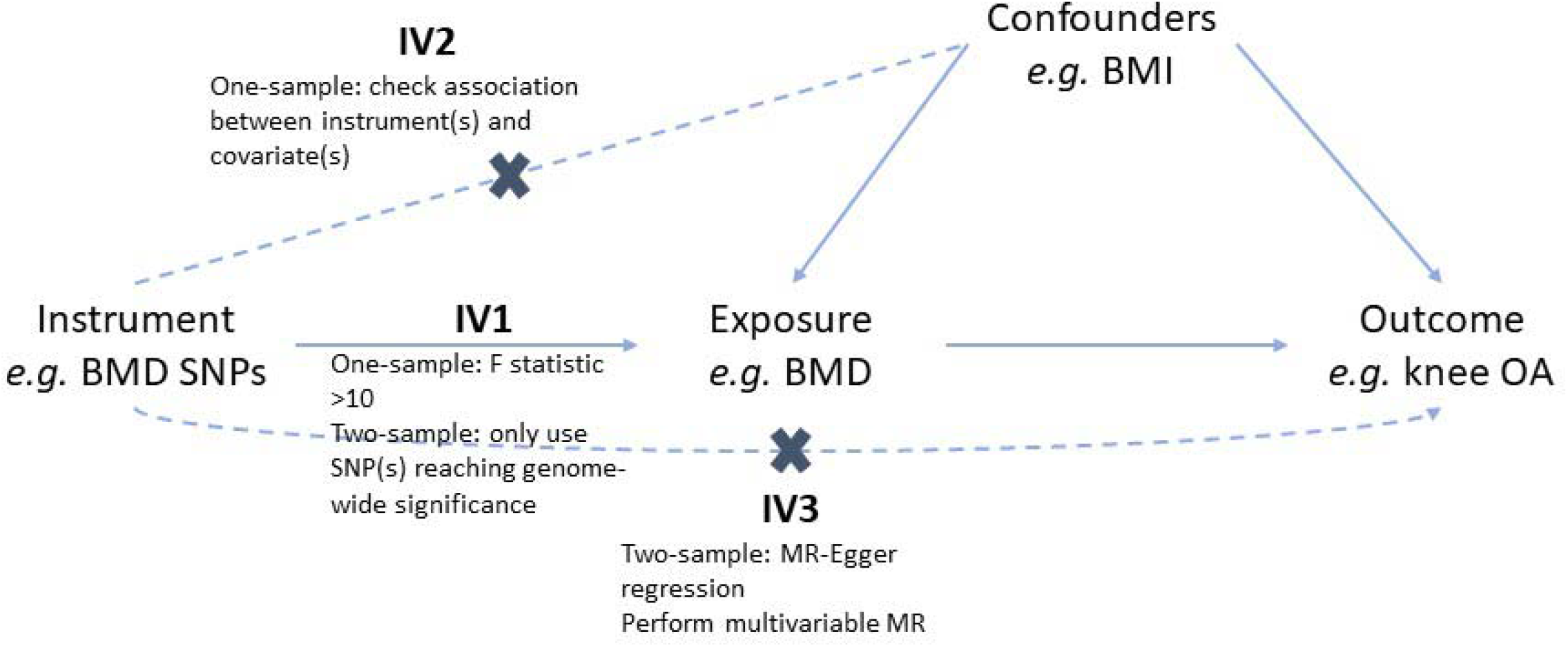
assumptions of Mendelian randomization and how we tested these assumptions. For an MR effect estimate to be valid, the instrument(s) must satisfy three key assumptions (32): IV1 (the instrument(s) must be robustly associated with the exposure); IV2 (the instrument(s) must not be associated with any confounders of the exposure-outcome relationship); and IV3 (the instruments(s) can only be associated with the outcome via the exposure and not via a different biological pathway independent of the exposure (i.e. horizontal pleiotropy). In one-sample analyses, IV1 was tested by calculating the F-statistic, which is a measure of instrument strength. A ≥10 threshold is used to determine sufficient instrument strength (2). IV2 was tested by determining the association between the instruments and potential confounders of the exposure-outcome relationship. In two-sample analyses, to satisfy IV1, we ensured that all instruments were robustly associated with the exposure by only including SNPs associated with the exposure at genome-wide significance. To address IV3, MR-Egger regression was performed to generate an estimate of horizontal pleiotropy (intercept) and a pleiotropy-robust estimate of the causal effect (slope). Weighted median regression was performed to determine the robustness of IVW estimates as weighted median estimates are valid even if up to 50% of the SNPs are not valid instruments (18). Abbreviations: BMD: bone mineral density; BMI: body mass index; OA: osteoarthritis; SNP: single-nucleotide polymorphism

#### One-sample MR

1SMR analyses were performed in the UKBB population using the instrumental-variable regression (‘ivreg’) function of the Applied Econometrics with R package (12). Exposures were instrumented by an unweighted genetic risk score (GRS), generated as the sum of the dosage for exposure-increasing alleles (data sources provided in *Table 1*). Analyses were adjusted for sex, genotyping chip and 10 principal components. Continuous exposures (eBMD/BMI) were standardized before analysis. Effect estimates for binary outcomes (hip/knee OA) were generated from a linear two-stage least-squares regression and represent the increased probability of having OA per unit increase in the exposure. We generated an estimate of the odds ratio per SD increase in the exposure, for comparison with 2SMR results, by first regressing the instruments on the exposure, generating predicted values of the exposure, and then regressing the predicted values of the exposure on the binary outcomes using a logistic regression model. The standard errors for this estimate are likely to be underestimated (13).

#### Two-sample MR

To maximise sample size, and thus statistical power, we performed 2SMR using summary-level data from published GWAS. 2SMR analyses were performed using the TwoSampleMR R package, version 0.4.22 (14). Single nucleotide polymorphism (SNP)-exposure estimates were extracted for all SNPs associated with the exposure at genome-wide significance. Details of the Genetic Factors for Osteoporosis (GEFOS), Genetic Investigation of Anthropometric Traits (GIANT) and the Genetics of Osteoarthritis (GO) consortia providing the summary statistics for eBMD, BMI and OA, respectively, are provided in the *Supplementary Information*. Summary statistics for the eBMD, BMI, hip and knee OA instruments are provided in *Supplementary Tables 2-7*. Clumping was performed to exclude non-independent SNPs based on a pairwise r^2^≥0.001. SNP-outcome effect estimates were then extracted for independent SNPs. SNP-outcome effect estimates for each analysis are presented in *Supplementary Tables 2-7*. SNP-exposure and SNP-outcome data were harmonized to ensure effect estimates corresponded to the same allele. Palindromic SNPs with indeterminate allele frequencies (MAF>0.42) were excluded. Steiger filtering excluded SNPs which explained a greater proportion of the variance in the outcome than the exposure (15): seven, four and two eBMD SNPs were excluded for analyses with hip OA, knee OA and BMI outcomes, respectively. Two BMI SNPs explained a greater proportion of variance in hip OA risk, one for knee OA risk and 15 for eBMD. One knee OA SNP was excluded due to a greater r^2^ for eBMD. All Steiger filtered SNPs are listed in *Supplementary Tables 2-7*. Estimates were generated using inverse-variance weighted (IVW) fixed-effects meta-analysis of the Wald ratios for each SNP.

#### Multivariable MR

As we hypothesized that BMI is a confounder of the BMD-OA relationship (*i*.*e*. a common causes of both phenotypes), we determined the independent effect of BMD on OA outcomes by performing 1S MVMR including GRS for both BMI and BMD as instruments. Both instruments were regressed on each exposure to generate a predicted value for each exposure. The predicted values for each exposure were then included in a multivariable regression to generate the effect of one exposure on OA when conditioning on the other exposure. Analyses were adjusted for sex, genotyping chip and 10 PCs. Sanderson-Windmeijer (SW) conditional F-statistics were calculated as measures of instrument strength in MVMR analyses (16).

#### Sensitivity analyses

MR-Egger regression was performed to generate an estimate of horizontal pleiotropy in the two-sample analyses (17). Weighted median regression determined the robustness of IVW estimates as weighted median estimates are valid even if up to 50% of the SNPs aren’t valid instruments (18). We repeated the 2SMR analyses restricted to eBMD SNPs also associated with FN-BMD (p≤5×10^−8^) in the GEFOS FN-BMD meta-analysis (19), to determine if FN-BMD has a stronger effect than eBMD on hip or knee OA risk. We also performed a latent causal variable model, as described by O’Connor and Price (20), to determine whether there is a true causal effect of BMD on OA, independent of the genetic correlation. Full methods are described in the *Supplementary Information*.

## Results

### Confirming observational relationships between BMD, OA and BMI in UKBB

334,061 individuals in UKBB with genotype data also had measurements of eBMD, BMI, covariates and hospital-diagnosed hip OA. 341,920 had data for knee OA. The mean(SD) age of those with hip OA was 61.7(6.0), of those with knee OA was 60.2(6.9) and of controls was 56.2(8.1) years (*Supplementary Table 8*). 57% of people with hip OA were female compared to 50% with knee OA and 54% of the controls. Both hip and knee OA cases were heavier than controls, with mean BMI 28.9(5.0), 30.3(5.4) and 27.1(4.6)kg/m^2^, respectively. Descriptive statistics were virtually the same when restricting to individuals with complete data for eBMD, BMI and OA who were included in the multivariable MR analyses (*Supplementary Table 8*).

In observational analyses adjusted for age and sex, an SD higher eBMD was associated with a 12% (95%CI:9,15) higher odds of hip OA and a 13% (11,15) higher odds of knee OA (*Supplementary Table 9*). A positive relationship was observed for eBMD and BMI, (standardized β=0.11[0.10,0.11]). An SD increase in BMI was associated with a 41% (39,44) higher odds of hip OA and a 73% (77,75) increased odds of knee OA.

### MR analyses provide evidence for bidirectional causal pathways between BMD and OA

A summary of MR results is presented in *Figure 3*. In 1SMR, eBMD was causally related to both hip and knee OA, with an SD increase in eBMD related to a 0.8% (0.1,1.4) increased probability of having hip OA and 1.7% (0.9,2.5) increased probability of having knee OA (*Table 2*). The F-statistic confirmed sufficient instrument strength (F>3000). The BMD risk score was related to BMI but wasn’t related to PA or HRT use (*Supplementary Table 10*). In 2SMR analyses, IVW provided evidence for a causal effect of eBMD on hip OA (OR per SD increase=1.09[1.03,1.16]), which was relatively consistent (in magnitude) across the three MR methods (*Figure 4, Supplementary Figure 1*). Evidence for a causal effect on knee OA was weaker (OR=1.04[1.00,1.09], *Supplementary Figure 2*). Excluding two SNPs more strongly related to BMI than eBMD did not alter results (*Supplementary Table 11*). When restricting to 10 SNPs also associated with FN-BMD (p≤5×10^−8^) in GEFOS, the magnitude of effect was stronger for hip OA (OR=1.40[1.12,1.74]), but this effect estimate was less consistent with the MR-Egger and weighted median estimates (*Supplementary Table 11*). However, evidence for a causal effect on knee OA, was more consistent (in magnitude) across the three methods (OR_IVW_=1.21[1.01,1.44]).

**Table 2:** results of one-sample univariable and multivariable MR

|  | Exposure | Outcome | N<sub>SNPs</sub> in GRS | F-statistic | N | Two-stage least-squares regression |  | Two-stage linear and logistic regression |  |
| --- | --- | --- | --- | --- | --- | --- | --- | --- | --- |
|  |  |  |  |  |  | estimate (95% CI) | p value | OR (95% CI) | p value |
| UV MR | eBMD | Hip OA | 44 | 3390 | 190 408 | 0.008 (0.001, 0.014)<sup>rd</sup> | 0.016 | 1.28 (1.05, 1.57) | 0.016 |
|  | eBMD | Knee OA | 44 | 3483 | 194 638 | 0.017 (0.009, 0.025)<sup>rd</sup> | 3x10<sup>-5</sup> | 1.40 (1.20, 1.63) | 3x10<sup>-5</sup> |
|  | BMI | Hip OA | 63 | 4201 | 333 828 | 0.015 (0.010, 0.021)<sup>rd</sup> | 6x10<sup>-8</sup> | 1.60 (1.35, 1.91) | 7x10<sup>-8</sup> |
|  | BMI | Knee OA | 63 | 4446 | 341 686 | 0.036 (0.029, 0.043)<sup>rd</sup> | <2x10<sup>-16</sup> | 2.01 (1.76, 2.29) | <2x10<sup>-16</sup> |
|  | BMI | eBMD | 63 | 2952 | 218 700 | 0.073 (0.037, 0.109)<sup>sd</sup> | 6x10<sup>-5</sup> |  |  |
|  | Hip OA | eBMD | 10 | 107 | 190 408 | 1.10 (0.36, 1.84)<sup>sd</sup> | 0.003 |  |  |
|  | Hip OA | BMI | 10 | 197 | 333 828 | 0.54 (0.01, 1.07)<sup>sd</sup> | 0.048 |  |  |
|  | Knee OA | eBMD | 4 | 49 | 194 638 | 4.16 (2.74, 5.57)<sup>sd</sup> | 8x10<sup>-9</sup> |  |  |
|  | Knee OA | BMI | 4 | 87 | 341 686 | 8.44 (6.65, 10.23)<sup>sd</sup> | <2x10<sup>-16</sup> |  |  |
| MV MR | eBMD | Hip OA | 44 | 275 | 190 175 | 0.007 (0.001, 0.013)<sup>rd</sup> | 0.027 | 1.26 (1.03, 1.54) | 0.027 |
|  | BMI | Hip OA | 63 | 211 | 190 175 | 0.013 (0.006, 0.020)<sup>rd</sup> | 6x10<sup>-4</sup> | 1.49 (1.19, 1.87) | 5x10<sup>-4</sup> |
|  | eBMD | Knee OA | 44 | 275 | 194 404 | 0.015 (0.008, 0.023)<sup>rd</sup> | 1x10<sup>-4</sup> | 1.36 (1.16, 1.59) | 1x10<sup>-4</sup> |
|  | BMI | Knee OA | 63 | 211 | 194 404 | 0.034 (0.025, 0.043)<sup>rd</sup> | 3x10<sup>-13</sup> | 1.93 (1.62, 2.30) | 3x10<sup>-13</sup> |
|  | Hip OA | eBMD | 10 | 7.5 | 190 175 | 1.12 (0.35, 1.89)<sup>sd</sup> | 0.004 |  |  |
|  | BMI | eBMD | 63 | 43 | 190 175 | 0.084 (0.040, 0.128)<sup>sd</sup> | 2x10<sup>-4</sup> |  |  |
|  | Knee OA | eBMD | 4 | 1.2 | 194 404 | 5.76 (2.62, 8.89)<sup>sd</sup> | 3x10<sup>-4</sup> |  |  |
|  | BMI | eBMD | 63 | 1.4 | 194 404 | -0.195 (-0.392, 0.001)<sup>sd</sup> | 0.052 |  |  |

<sup>sd</sup> estimate represents the SD increase in outcome per SD increase in exposure. Estimates for binary exposures represent the SD increase in outcome per doubling in risk of exposure.
<sup>rd</sup> Estimates for binary outcomes represent the risk difference per SD increase in exposure. Due to the difficult interpretation of these results, odds ratios were also calculated by two-stage regression. Odds ratios are per SD increase in exposure.
Abbreviations: MR: mendelian randomization; BMD: bone mineral density; OA: osteoarthritis; UV: univariable; MV: multivariable; eBMD: estimated BMD

**Figure 3:**
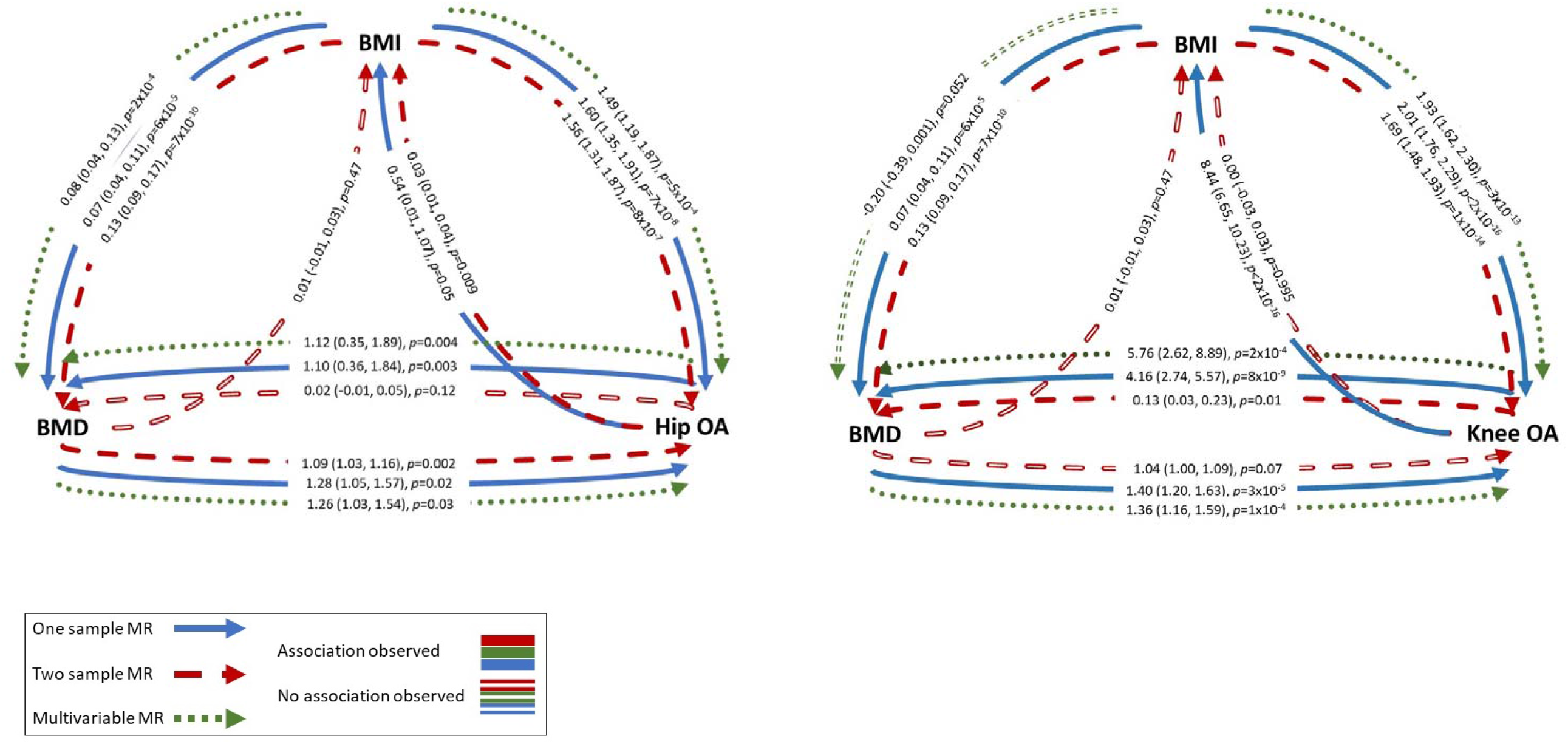
summary of results of one-sample, two-sample and multivariable MR analyses. Effect estimates represent the SD increase in outcome per SD increase in exposure for BMD-BMI and BMI-BMD analyses, the odds ratio per SD increase in exposure for BMI-OA and BMD-OA analyses and the SD increase in BMD or BMI per 1 unit increase in the log odds of OA Abbreviations: BMD: bone mineral density; BMI: body mass index, OA: osteoarthritis; MR: Mendelian randomization

**Figure 4:**
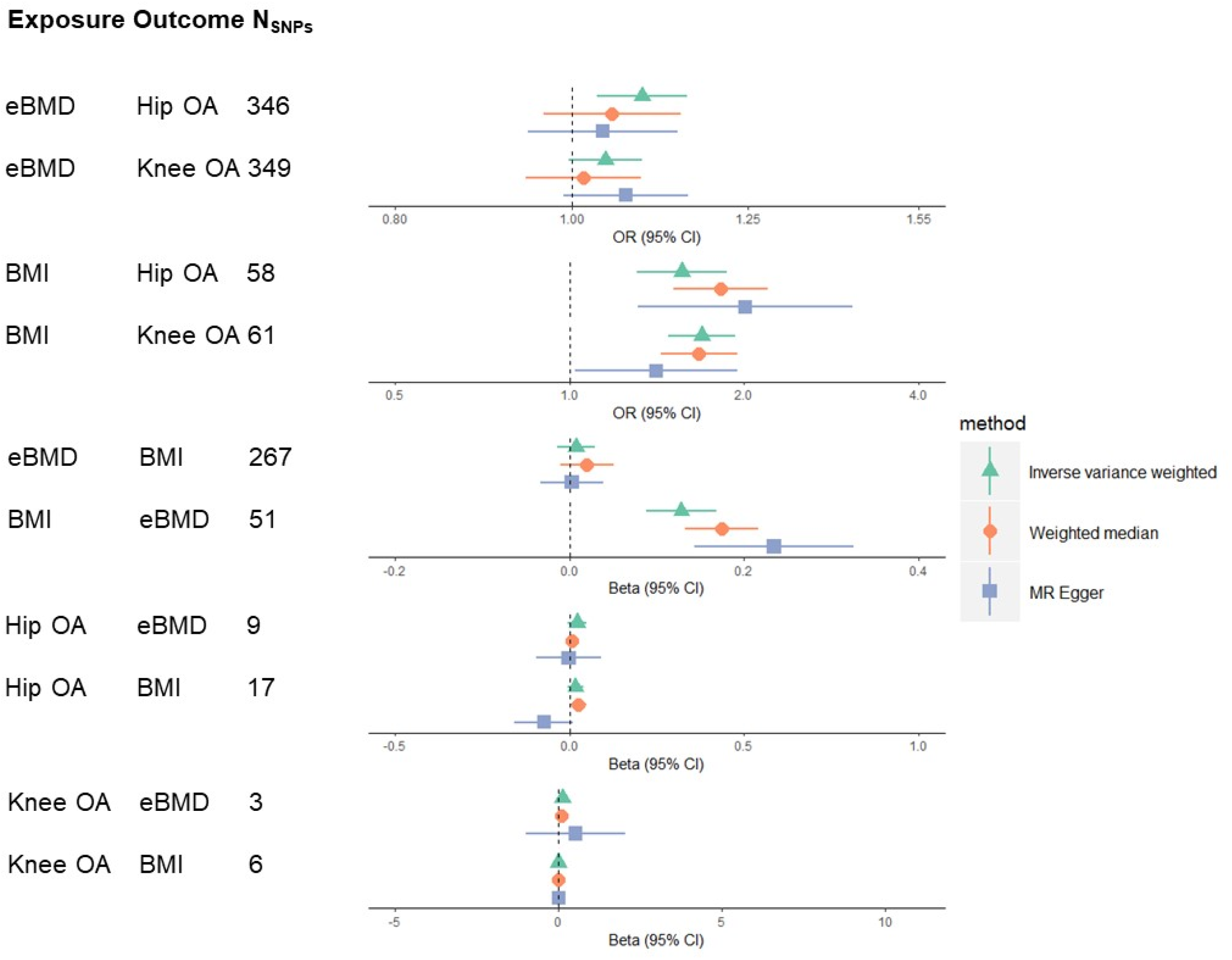
results of two-sample MR analyses. Abbreviations: eBMD: estimated bone mineral density; OA: osteoarthritis; BMI: body mass index; CI: confidence interval

There was evidence for a causal pathway between hip OA and eBMD in 1SMR (SD increase per doubling in odds of hip OA=1.10[0.36,1.84]) (*Table 2*), but not 2SMR analysis (*Supplementary Figure 3*). Evidence for a causal effect of knee OA on eBMD was provided by 1SMR (β=4.16[2.74,5.57]) and 2SMR (β=0.13[0.03,0.23]), with a positive effect observed for all three 2SMR methods, albeit weaker, with wide CIs overlapping the null for MR-Egger regression (*Figure 4, Supplementary Figure 4*). The knee, but not hip, OA GRS was related to BMI, potentially invalidating IV2 (*Supplementary Table 10*).

#### BMI is a strong causal risk factor for BMD and OA with weaker evidence for bidirectionality

1SMR provided evidence that BMI has a strong causal effect on hip and knee OA, with an SD increase in BMI associated with a 1.5% (1.0,2.1) increased risk of hip OA and 3.6% (2.9,4.3) increased risk of knee OA (*Table 2*). 2SMR suggested that BMI is causally related to OA, with an SD increase in BMI related to a 56% (31,87) increased odds of hip OA and a 69% (48,93) increased odds of knee OA. These results were consistent across the three 2SMR methods (*Supplementary Figures 5,6*), other than the causal effect of BMI on knee OA estimated by MR Egger, which was approximately 30% weaker, albeit in the same direction (*Figure 4*). There was strong evidence, from 1SMR, that the causal pathway between BMI and knee OA was bidirectional, with weaker evidence for hip OA (*Table 2*). Additional adjustment for total weekly PA (assessed using the IPAQ questionnaire) did not attenuate these relationships. 2SMR, however, provided weak and inconsistent evidence (across the three methods) of a causal effect of hip OA on BMI only (*Figure 4, Supplementary Figures 7, 8*).

We could not perform bidirectional 1SMR for BMD-BMI as the FN-BMD SNPs were identified by weight-adjusted GWAS, meaning the instrument for FN-BMD may be inversely related to weight and thus BMI (21). 2SMR using summary statistics from the eBMD GWAS, not adjusted for weight, did not identify a causal effect of eBMD on BMI (*Figure 4, Supplementary Figure 9*). There was robust evidence for a causal effect of BMI on eBMD in 1SMR, with an SD increase in BMI causing a 0.07SD (0.04,0.11) increase in heel BMD (*Table 2*). This estimate was like that from 2SMR and the effect size was consistent for IVW, weighted median and MR Egger analyses, although the MR-Egger intercept did reveal some evidence of horizontal pleiotropy (*Figure 4, Supplementary Table 11, Supplementary Figure 10*).

### Multivariable MR identifies an independent causal effect of eBMD on OA

Overall, the 1S and 2S analyses provided consistent evidence that BMI is a confounder of the relationship between BMD and hip/knee OA (*i*.*e*. a common cause of both phenotypes, *Figure 3*). We therefore used 1SMVMR to examine the causal effect of eBMD on OA after accounting for BMI. Following adjustment for BMI, eBMD was found to be an independent causal risk factor for both hip and knee OA with a similar magnitude of effect to that observed in MR analyses not accounting for BMI. BMI had a stronger effect than eBMD for both hip and knee OA (*Table 2*). SW F-statistics were >200 for both instruments.

MVMR provided evidence for a BMI-independent causal effect of OA on eBMD (β_hip_=1.12[0.35,1.89], β_knee_=5.76[2.62,8.89], *Table 2*). The causal effect of BMI on BMD was independent of hip OA (β=0.08[0.04,0.13]). When conditioning on knee OA, an inverse effect of BMI on BMD was observed (β=-0.20[-0.39,0.001]). This is unlikely to be bias due to conditioning on a common outcome (*i*.*e*. collider bias), as genetically-predicted OA is not a common outcome (16). However, of note these results are vulnerable to weak instrument bias as conditional F statistics were <10.

### Latent causal variable analyses provide evidence for a non-pleiotropic causal effect of BMD on OA

To determine if shared underlying genetic aetiology fully explained the observed causal effect of BMD on OA, we performed latent causal variable modelling using weight-adjusted summary statistics for both FN/LS BMD and hip/knee OA. The LCV analysis identified evidence for genetic correlations between BMD (measured at both the FN and LS) and OA at both the hip and knee (rho=0.16-0.23, *Supplementary Table 12*). There was evidence for a partial causal effect of BMD at both sites on OA at both sites, independent of genetic correlation and weight, with the largest magnitude of causal effect observed for FN BMD and knee OA, with a genetic causality proportion of 0.64.

## Discussion

We have found strong evidence for a causal effect of BMD on hip and knee OA using 1SMR, which was relatively consistent with 2SMR. MVMR confirmed that the effect of BMD on OA is independent of BMI. Our results also suggest that there is a bidirectional causal effect between OA and eBMD. We have confirmed strong causal effects of BMI on eBMD, hip and knee OA, with no causal effect of eBMD on BMI. Finally, we have found some evidence of a positive causal effect of knee OA on BMI. The observed causal effect of BMI on eBMD in this adult population is consistent with a previous analysis of a paediatric population (mean age 10), where a causal effect of BMI on FN-BMD was observed (8). As seen in this current analysis, Kemp *et al* found no evidence for a causal effect of BMD on BMI (8). The strong causal effect of BMI on both hip and knee OA corroborates previous MR analyses (4, 22).

The causal effect of eBMD on hip and knee OA which we observed is consistent with previous MR analyses identifying causal effects of FN and LS-BMD on hip and knee OA (4, 22). Taken together, these findings suggest that bone parameters in general have a causal effect on OA, regardless of the site or method of measurement. However, the magnitude of effect of eBMD on OA was larger in 2S analyses restricted to SNPs associated with FN-BMD. There are two potential explanations for a stronger effect of BMD on OA when restricting to FN-BMD loci. Firstly, FN-BMD measured by DXA may be a more accurate representation of the biological pathways between bone and cartilage, compared to eBMD, which represents a combination of speed of sound and broadband ultrasound attenuation. Alternatively, since the FN primarily comprises cortical bone, whereas heel BMD is predominantly trabecular (23, 24), these findings may reflect the fact that cortical bone is more strongly related to OA pathogenesis compared to trabecular bone. For example, cortical BMD might be expected to correlate more strongly with subchondral plate sclerosis, compared to trabecular BMD, which is implicated in the progression of OA (25).

We have found some evidence for reverse causality in the relationship between eBMD and OA. The positive direction of effect is as expected from artefactual elevation, rather than loss of bone mass due to reduced PA. However, as we do not expect BMD measurements at the heel to be artefactually elevated by features of OA, the observed causal effect of OA on eBMD in 1SMR may reflect biological pleiotropy (*i*.*e*. the same underlying biological pathways may be contributing to both phenotypes). Consistent with shared biological mechanisms contributing to both BMD and OA, Hackinger *et al* identified a genetic correlation between LS-BMD (but not FN) and OA (10). By performing a cross-phenotype meta-analysis between OA and LS-BMD, the authors identified a number of known loci, as well as a novel locus in the SMAD3 gene (10). SMAD3 is part of the transforming growth factor β (TGFβ) signalling pathway, which regulates osteoblast differentiation. The first discovered OA loci, growth differentiation factor-5 (GDF5), is a ligand for this pathway (26). The canonical Wnt signalling pathway is involved in the regulation of osteoblasts and mutations in this pathway can lead to high or low BMD; for example activating mutations in low-density lipoprotein receptor-related protein 5 (LRP5, the receptor involved in Wnt signalling activation) cause high BMD (46). This signalling pathway has been implicated in OA pathogenesis (47); increased levels of a Wnt signalling inhibitor, DKK1, were associated with reduced progression of hip OA in a population of Caucasian women (48).

However, we did find stronger, more consistent, evidence for an effect of eBMD on OA, as opposed to vice versa. This could reflect the stronger instrument for BMD, but our LCV analyses using the full set of summary statistics provided further evidence for a causal pathway between BMD and OA, independent of genetic correlation (and confounding by weight), suggesting that bone may still have a direct effect on OA, for example via increased joint loading, or structural differences in the subchondral bone, which have been linked to progression of joint space narrowing (JSN) (27).

### Strengths and limitations

We have utilised the largest datasets possible to maximize the power to detect causal effects. We have ensured that there is no overlap between our exposure and outcome populations. We have examined individual-level data in UKBB to perform 1SMR to strengthen evidence.

However, we were unable to use eBMD instruments for 1SMR as they were identified in the same population used for analysis; reassuringly F-statistics suggested that our FN-BMD instrument was of reasonable strength. Our OA outcomes for 1SMR were based on hospital-diagnosis; it is unclear how this phenotype relates to radiographic features of OA, such as JSN, which are commonly used as clinical trial outcomes. Using a severe phenotype as the outcome means reduced power in GWAS and leads to fewer genome-wide significant loci and a greater chance of weak instrument bias (as highlighted by the much smaller F-statistics for the OA instruments). The OA outcomes from the GO consortium included a range of definitions of hip and knee OA, including hospital diagnosis, radiographic evidence and self-reported OA definitions. Heterogeneity in phenotype also reduces the power to detect loci in GWAS. The ORs from 1SMR are estimates and SEs are likely underestimated (13), therefore caution should be taken when interpreting these effect sizes. There may be additional risk factors related to the genetic variants which we did not include in our MVMR models. All populations were of white European ancestry, limiting generalizability to other ethnicities.

### Conclusions

We have found evidence for a BMI-independent causal effect of BMD on hip and knee OA and some evidence for a bidirectional causal effect, which we hypothesize to reflect shared underlying genetic aetiology. We have confirmed strong causal effects of BMI on BMD and hip and knee OA and have found novel evidence for a causal effect of knee OA on BMI, which did not appear to be mediated by pain-associated reductions in physical activity. Further analyses are required to determine the shared pathways contributing to both BMD and OA, and to determine the mechanisms by which higher BMD causes OA.

## Data Availability

UK Biobank data are available through a procedure described at https://www.ukbiobank.ac.uk/principles-of-access/. BMD GWAS summary statistics are available to download from the GEFOS website at http://www.gefos.org/?q=content/gefos-data-release. GIANT BMI GWAS summary statistics are available to download at https://portals.broadinstitute.org/collaboration/giant/index.php/GIANT_consortium_data_files#GIANT_Consortium_2012-2015_GWAS_Summary_Statistics. Hip and knee OA GWAS summary statistics from UK Biobank and arcOGEN are publicly available at https://www.ebi.ac.uk/gwas/. 

## Acknowledgements

This work was supported by the Wellcome Trust (grant ref 20378/Z/16/Z). CLG was funded by Versus Arthritis (grant ref 20000). JZ receives salary and start-up funding from the University of Bristol (Vice-Chancellor’s fellowship). JZ is also supported by the Academy of Medical Sciences (AMS) Springboard Award, the Wellcome Trust, the Government Department of Business, Energy and Industrial Strategy (BEIS), the British Heart Foundation and Diabetes UK (SBF006\1117). AH, ES, RG, LP, JZ, GDS, CLG and JHT work in, or are affiliated with, a University of Bristol and MRC funded unit (MC_UU_00011/1, MC_UU_00011/4). Individual-level analyses have been conducted using the UK Biobank Resource under Application Number 17295.

