## Supplementary Information for "Using multivariable Mendelian randomization to estimate the causal effect of bone mineral density on osteoarthritis risk, independent of body mass index"

### The Genetics of Osteoarthritis consortium

#### Names and affiliations of consortium members not list in the main author list

Lilja Stefánsdóttir^1^, Yanfei Zhang^2^, Rodrigo Coutinho de Almeida^3^, Tian T. Wu^4^, Jie Zheng^5^, Maris Teder-Laving^6^, Anne-Heidi Skogholt^7^, Chikashi Terao^8^, Eleni Zengini^9^, George Alexiadis^10^, Andrei Barysenka^11^, Gyda Bjornsdottir^1^, Maiken E. Gabrielsen^7^, Arthur Gilly^11^, Thorvaldur Ingvarsson^12,13^, Marianne B. Johnsen^7,14,15^, Helgi Jonsson^12,16^, Margreet G. Kloppenburg^17^, Almut Luetge^7^, Reedik Mägi^6^, Massimo Mangino^18^, Rob R.G.H.H. Nelissen^19^, Manu Shivakumar^20^, Julia Steinberg^11,21,22,23^, Hiroshi Takuwa^24,25^, Laurent Thomas^26,7,27,28^, Margo Tuerlings^1^, George Babis^29^, Jason Pui Yin Cheung^30^, Dino Samartzis^30^, Steve A. Lietman^31^, P. Eline Slagboom^3^, Kari Stefansson^3,12^, André G. Uitterlinden^32^, Bendik Winsvold^7,33,34^, John-Anker Zwart^7,33^, Pak Chung Sham^35^, Gudmar Thorleifsson^1^, Tom R Gaunt^5^, Andrew P. Morris^36^, Ana M. Valdes^37^, Aspasia Tsezou^38^, Kathryn S.E Cheah^39^, Shiro Ikegawa^24^, Kristian Hveem^7,40^, Tõnu Esko^6^, J Mark Wilkinson^41^, Ingrid Meulenbelt^3^, Ming Ta Michael Lee^2,42^, Unnur Styrkársdóttir^1^

^1^ deCODE genetics / Amgen Inc., Reykjavik, Iceland

^2^Genomic Medicine Institute, Geisinger Health System, Danville, PA 17822, USA

^3^ Department of Biomedical Data Sciences, Section Molecular Epidemiology, Leiden University Medical Center, Leiden, The Netherlands

^4^Department of Psychiatry, Li Ka Shing Faculty of Medicine, The University of Hong Kong, Hong Kong

^5^MRC Integrative Epidemiology Unit (IEU), Bristol Medical School, University of Bristol, Oakfield House, Oakfield Grove, Bristol, BS8 2BN, United Kingdom

^6^Estonian Genome Center, Institute of Genomics, University of Tartu, Tartu, Estonia

^7^K. G. Jebsen Center for Genetic Epidemiology, Department of Public Health and Nursing, Faculty of Medicine and Health Sciences, Norwegian University of Science and Technology, Trondheim, Norway

^8^Laboratory for Statistical and Translational Genetics, RIKEN Center for Integrative Medical Sciences, Kanagawa, Japan

^9^5th Psychiatric Department, Dromokaiteio Psychiatric Hospital, Haidari, Athens, Greece

^10^1^st^ Department of Orthopaedics, KAT General Hospital, Athens, Greece

^11^ Institute of Translational Genomics, Helmholtz Zentrum München, German Research Center for Environmental Health, Neuherberg, Germany

^12^Faculty of Medicine, University of Iceland, Reykjavik, Iceland

^13^Department of Orthopedic Surgery, Akureyri Hospital, Akureyri, Iceland

^14^Research and Communication Unit for Musculoskeletal Health (FORMI), Department of Research, Innovation and Education, Division of Clinical Neuroscience, Oslo University Hospital, Oslo, Norway

^15^Institute of Clinical Medicine, Faculty of Medicine, University of Oslo, Oslo, Norway

^16^Department of Medicine, Landspitali The National University Hospital of Iceland, Reykjavik, Iceland

^17^Department of Rheumatology, Leiden University Medical Center, Leiden, The Netherlands

^18^Department of Twin Research and Genetic Epidemiology, Kings College London, London, United Kingdom

^19^Department of Orthopaedics, Leiden University Medical Center, Leiden, The Netherlands

^20^Department of Biostatistics, Epidemiology and Informatics, Perelman School of Medicine, University of Pennsylvania, Philadelphia, PA, USA

^21^Cancer Research Division, Cancer Council NSW, Sydney, NSW, Australia

^22^School of Public Health, Faculty of Medicine and Health, The University of Sydney, Australia

^23^Human Genetics, Wellcome Genome Campus, Wellcome Sanger Institute, Cambridge, United Kingdom

^24^Laboratory for Bone and Joint Diseases, RIKEN Center for Integrative Medical Sciences, Tokyo, Japan

^25^Department of Orthopedic Surgery, Shimane University, Izumo, Japan

^26^Department of Clinical and Molecular Medicine, Norwegian University of Science and Technology, Trondheim, Norway

^27^BioCore - Bioinformatics Core Facility, Norwegian University of Science and Technology, Trondheim, Norway

^28^Clinic of Laboratory Medicine, St. Olavs Hospital, Trondheim University Hospital, Trondheim, Norway

^29^ 2nd Department of Orthopaedics, National and Kapodistrian University of Athens, Medical School, Nea Ionia General Hospital, "Konstantopouleio", Athens, Greece

^30^Department of Orthopaedics and Traumatology, The University of Hong Kong, Hong Kong

^31^Musculoskeletal Institute, Geisinger, Danville, USA

^32^Department of Internal Medicine, Erasmus MC, Medical Center, Rotterdam, The Netherlands

^33^Department of Research, Innovation and Education, Division of Clinical Neuroscience, Oslo University Hospital, Oslo, Norway

^34^Department of Neurology, Oslo University Hospital, Oslo, Norway

^35^Li Ka Shing Faculty of Medicine, The University of Hong Kong, Hong Kong

^36^Centre for Genetics and Genomics Versus Arthritis, Centre for Musculoskeletal Research, University of Manchester, Manchester, United Kingdom

^37^Faculty of Medicine & Health Sciences, School of Medicine, University of Nottingham, Nottingham, Nottinghamshire, United Kingdom

^38^Laboratory of Cytogenetics and Molecular Genetics, Faculty of Medicine, University of Thessaly, Larissa, Greece

^39^School of Biomedical Sciences, The University of Hong Kong, Hong Kong

^40^HUNT Research Center, Department of Public Health and Nursing, Faculty of Medicine and Health Sciences, Norwegian University of Science and Technology, Trondheim, Norway

^41^Department of Oncology & Metabolism and Healthy Lifespan Institute, University of Sheffield, Sheffield, United Kingdom

^42^Institute of Biomedical Sciences, academia Sinica, Taipei, Taiwan

### Supplementary methods

#### UK Biobank population

UKBB is a UK-wide population-based health research resource consisting of approximately 500,000 people, aged 38-73, who were recruited 2006-2010 (1). Participants provided a range of information (e.g. demographics, health status, lifestyle/PA measures) via questionnaires and interviews; anthropometric measures and blood samples were taken (data available at [www.ukbiobank.ac.uk](http://www.ukbiobank.ac.uk)). A full description of the study design, participants and quality control (QC) methods has been published (1). Methods for assessing BMD and ascertaining hospital-diagnosed OA status are described in the *Supplementary Information.* UKBB received ethical approval from the Research Ethics Committee (REC reference:11/NW/0382).

##### eBMD measurement

BMD was estimated from ultrasound measurement of the calcaneus using a Sahara Clinical Bone Sonometer. eBMD is estimated from a combination of speed of sound and broadband ultrasound attenuation (2).

##### Hospital-diagnosed OA ascertainment

Hospital-diagnosed hip and knee OA were determined from hospital episode statistics (HES) (3) using International Statistical Classification of Diseases and Related Health Problems (ICD) 9/10 codes previously reported for knee and hip OA (4). Inclusion codes for cases and exclusion codes for controls (*e.g.* to exclude controls with OA in other joints) are included as *Supplementary Table 1A* and *Supplementary Table 1B,* respectively.

##### Genotyping and imputation

The UKBB full data release contains data on all successfully genotyped samples (n=488,377). 49,979 individuals were genotyped using the UK BiLEVE array and 438,398 using the UKBB axiom array. Pre-imputation QC, phasing and imputation are described elsewhere (5), as well as QC filtering steps (6). Briefly, prior to phasing, multiallelic and rare SNPs (minor allele frequency [MAF] ≤1%) were removed (5). Phasing of genotype data was performed using a modified version of the SHAPEIT2 algorithm (5). Genotype imputation to a reference set, combining the UK10K haplotype and HRC reference panels (7), was performed using IMPUTE2 algorithms (8). Analyses were restricted to autosomal variants by graded filtering with varying imputation quality according to allele frequency ranges. Therefore, rarer genetic variants are required to have a higher imputation info score (info>0.3 for MAF >3%; info>0.6 for MAF 1-3%; info>0.8 for MAF 0.5-1%; info>0.9 for MAF 0.1-0.5%) with MAF and info scores having been recalculated on an in-house derived ‘European’ subset.

QC filtering of the UK Biobank (UKBB) data was performed by Mitchell *et al* as described in the published protocol (6). In brief, individuals with mismatches between genetic and reported sex or individuals with sex chromosome aneuploidy were excluded. The sample was restricted to individuals of European ancestry as defined by an in-house k-means cluster analysis performed using the first 4 principal components (PCs) provided by UKBB in the statistical software environment R. The current analysis uses the largest cluster from this analysis. Estimated kinship coefficients, using the KING toolset (9), identified pairs of related individuals (5). An in-house algorithm was applied to preferentially remove individuals related to the greatest number of other individuals, until no related pairs remained.

##### Observational analysis

Observational associations with BMI or eBMD as the outcome were determined by multivariable linear regression. Analyses with OA as the outcome were performed using multivariable logistic regression. All analyses were adjusted for age at baseline assessment and sex.

#### 2SMR data sources

##### GEFOS

The Genetic Factors for Osteoporosis (GEFOS) consortium GWAS is a collaboration of 17 discovery cohorts (n=32 961) from North America, Europe, East Asia and Australia and 34 replication populations (n=50 933) which aimed to identify genetic variants associated with FN-BMD and LS-BMD (10). Full methodology has been published elsewhere (10). This meta-analysis represents the largest GWAS to-date of FN-BMD and therefore FN-BMD genome-wide significant loci from this analysis were selected to instrument eBMD in UKBB (as an eBMD instrument not generated in UKBB does not exist). More recently, the GEFOS consortium have performed a GWAS of eBMD including 426,824 individuals from UKBB, which represents the largest eBMD GWAS to date and provided the summary statistics for 2SMR analyses with eBMD (11). This GWAS was adjusted for age, sex, genotyping array, assessment centre and PCs.

##### GIANT

BMI summary statistics for 2S analyses were taken from the two-stage GWAS meta-analysis performed by the Genetic Investigation of Anthropometric Traits (GIANT) consortium (12). This meta-analysis was performed in two stages: stage one included 80 BMI GWAS (n=234 069) and stage two included an additional 34 GWAS (n=88 137) (12). The GIANT meta-analysis represents the largest BMI GWAS which does not include the UKBB population and we could therefore use the summary statistics for 2S analyses with UKBB-derived outcome summary statistics and to instrument BMI in UKBB. The GIANT BMI GWAS was adjusted for age, age^2^ and PCs.

##### GO consortium

The Genetics of Osteoarthritis (GO) consortium is a collaboration of cohorts aiming to identify novel genetic variants for OA. The most recent published GO GWAS meta-analysed GWAS from the UKBB and arcOGEN populations and included 77 052 cases and 378 169 controls, adjusting for age, sex, genotyping chip and PCs (4). ArcOGEN is a population of UK-based Europeans with clinically-diagnosed knee and/or hip OA (13). More recently, the GO consortium has expanded to include additional cohorts and the total sample size is now >800,000 from 13 international cohorts, with full details of cohort-specific GWAS published elsewhere (14). This has allowed the largest GWAS meta-analysis for OA excluding UKBB individuals to be performed, generating summary statistics which could be used for 2S analyses when the SNP-exposure summary statistics have been generated in the UKBB population.

#### Latent causal variable model

The Latent Causal Variable (LCV) model has been developed by O’Connor and Price (15) to estimate the causal effect of one trait on another trait, when the two traits are genetically correlated. The genetic correlation between the two traits is modelled as a latent variable, and the genetic correlation between the latent variable and each trait is assessed. A perfect genetic correlation between the latent variable and one of the traits suggests a fully genetically causal effect of that trait on the other trait. If the trait has a partial causal effect on the (outcome) trait of interest (*i.e.* the genetic correlation between the latent variable and the (exposure) trait is less than 1), partial causality can be estimated using the genetic causality proportion (GCP), with a value of 0 representing no causal effect and a value closer to 1 representing a stronger causal effect.

Three datasets are required to run this model: genome-wide summary statistics for SNP-BMD associations, genome-wide summary statistics for SNP-OA associations and LD scores for each SNP. The LD scores were generated from the 1000 Genomes European population and were download from ‘<https://data.broadinstitute.org/alkesgroup/LDSCORE/>’ (eur_w_ld_chr.tar.bz). To maximise sample size for the OA summary statistics whilst limiting to a fully-European population, we aimed to use OA GWAS summary statistics generated from UK Biobank. We therefore could not use the eBMD summary statistics for the SNP-exposure relationships and therefore used the Estrada summary statistics, adjusted for weight. We performed GWAS of both hospital-diagnosed hip and knee OA, adjusting for weight, within UKBB. GWAS was performed as described in the published protocol (16), adjusting for sex, genotyping chip and 10 PCs, as well as weight measured at the assessment clinic. 410 052 individuals were included in the GWAS of knee OA and 400 516 in the GWAS of hip OA. QQ plots are provided in *Supplementary Figure 11.*

The MHC region (Chr 6, 28.5-33.5Mb) was removed from both sets of summary statistics, as well as SNPs with a MAF<0.05. Datasets were restricted to SNPs present in both datasets. Alleles were harmonized so that the beta corresponded to the same effect allele. Betas were then transformed by dividing them by their SE. Analyses were then performed using the ‘RunLCV.R’ script provided by the authors (<https://github.com/lukejoconnor/LCV>).

#### Supplementary Tables

###### Supplementary Table 1A: ICD codes used to identify cases of hospital-diagnosed hip and knee OA

| **Joint** | **Code** | | **Description** |
| --- | --- | --- | --- |
|  | **ICD10** | **ICD9** |  |
| Hip | M16 |  | “Coxarthrosis” |
|  | M160 |  | “Primary coxarthrosis, bilateral” |
|  | M161 |  | “Other primary coxarthrosis” |
|  | M169 |  | “Coxarthrosis, unspecified” |
|  | M1905 |  | “Primary arthrosis of other joints (pelvic region and thigh)” |
|  | M1995 |  | “Arthrosis, unspecified (pelvis region and thigh)” |
|  |  | 71535 | “Osteoarthrosis, localized, primary or secondary, pelvic region and thigh” |
|  |  | 71515 | “Osteoarthrosis, localized, primary, pelvic region and thigh” |
| Knee | M17 |  | “Gonarthrosis” |
|  | M170 |  | “Primary gonarthrosis, bilateral” |
|  | M171 |  | “Other primary gonarthrosis” |
|  | M179 |  | “Gonarthrosis, unspecified” |
|  | M1906 |  | “Primary arthrosis of other joints (lower leg)” |
|  | M1996 |  | “Arthrosis, unspecified (lower leg)” |
|  |  | 71536 | “Osteoarthrosis, localized, primary or secondary, lower leg” |
|  |  | 71516 | “Osteoarthrosis, localized, primary, lower leg” |

*Abbreviations: ICD: International Statistical Classification of Diseases and Related Health Problems*

###### Supplementary Table 1B: ICD codes used to exclude controls with OA at other sites or other arthropathies for hospital-diagnosed hip, knee and hand OA

|  |  | **ICD10** | | | | | | | **ICD9** | | | | |
| --- | --- | --- | --- | --- | --- | --- | --- | --- | --- | --- | --- | --- | --- |
| M111*  M112*  M118*  M119*  M13  M130  M1300  M1305  M1306  M1309  M131  M1310  M1314  M1315  M1316  M1319  M138  M1380  M1384  M1385  M1386  M1389  M139  M1390  M1394  M1395  M1396  M1399  M15  M150  M151 | M152  M153  M154  M158  M159  M1599  M16  M160  M161  M162  M163  M164  M165  M166  M167  M169  M17  M170  M171  M172  M173  M174  M175  M179  M18  M180  M181  M182  M183  M184  M185 | M189  M19  M190*  M191*  M192*  M198*  M199*  M20  M200  M210  M2100  M2105  M2106  M2109  M211  M2110  M2115  M2116  M2119  M212  M2120  M2124  M2125  M2126  M2129  M22  M220  M221  M222  M223  M224 | M228  M229  M23  M230  M2300  M2301  M2302  M2303  M2304  M2305  M2306  M2307  M2309  M231  M2310  M2311  M2312  M2314  M2315  M2316  M2317  M2319  M232  M2320  M2321  M2322  M2323  M2324  M2325  M2326  M2327 | M2329  M233  M2330  M2331  M2332  M2333  M2334  M2335  M2336  M2337  M2339  M234  M2340  M2341  M2342  M2343  M2344  M2345  M2346  M2347  M2349  M235  M2350  M2351  M2352  M2353  M2354  M2355  M2356  M2357  M2359 | M236  M2360  M2361  M2362  M2363  M2364  M2365  M2366  M2367  M2369  M238  M2380  M2381  M2382  M2383  M2384  M2385  M2386  M2387  M2389  M239  M2390  M2391  M2392  M2393  M2394  M2395  M2396  M2397  M2399  M24 | M240*  M241*  M242*  M243  M2430  M2434  M2435  M2436  M2439  M244  M2440  M2444  M2445  M2446  M2449  M245  M2450  M2454  M2455  M2456  M2459  M246  M2460  M2464  M2465  M2466  M2469  M247*  M248  M2480  M2484 | M2485  M2486  M2489  M249  M2490  M2494  M2495  M2496  M2499  M25  M255*  M256*  M257*  M258*  M259*  M42  M420*  M421*  M429*  M472*  M478*  M479  M4790  M4791  M4792  M4793  M4794  M4795  M4796  M4797  M4798 | M4952  M4953  M4954  M4955  M4956  M4957  M4958  M4959  M498*  M50  M501  M502  M503  M508  M509  M51  M949*  Q65*  V134  V135  V136  M21 | | 712  7121  7122  7123  7128*  7129*  715  7150  7151*  7152*  7153*  7158  7159  7161*  7165  71650  71654  71655  71656  71659  7166  71660  71664  71665  71666  71669  7168*  7169*  717  7170  7171 | 7172  7173  7174  7175  7176  7178  7179  718  7180*  7181*  7182*  7183*  7184*  7185*  7186*  7188*  7189*  7192*  7194*  7195*  7196*  7197  7198*  7199*  7200  721*  722  7220  7221  7222  7223 | 7224  7225  7226  7229  723  7230  7231  7295*  73100  73101  73102  73103  73104  732  7320  7321  7322  7323  7324  7325  7326  7327*  7328*  7329  7339*  736  7362  7363  7364  7365  7366 | 7367  7368  7369  7543  75430  75431  75432  7544  75440  75441  75442  75443  75444  7545  75450  75451  75452  75453  75459  7546  75460  75461  75469  835  8350  8351  836  8360  8361  8362  8366 |

**represents any number from 1 to 9*

###### Supplementary Tables 2 and 3: see separate excel file

###### Supplementary Table 4: Associations between the SNPs used to instrument hip OA in 2SMR and hip OA in the GO consortium and eBMD in UK Biobank

|  | | **Exposure=Hip OA** | | | | **Outcome=eBMD** | | | |  |
| --- | --- | --- | --- | --- | --- | --- | --- | --- | --- | --- |
| **SNP** | **EA** | **EAF** | **Beta** | **SE** | **P** | **EAF** | **Beta** | **SE** | **p** | **passed Steiger filtering^a^** |
| rs10843013 | A | 0.781 | -0.105 | 0.014 | 8.40x10^-15^ | 0.794 | 0.000 | 0.002 | 8.50x10^-1^ | TRUE |
| rs11164653 | T | 0.418 | -0.076 | 0.011 | 2.46x10^-11^ | 0.409 | -0.004 | 0.002 | 2.40x10^-2^ | TRUE |
| rs12209223 | A | 0.116 | 0.125 | 0.018 | 8.76x10^-13^ | 0.102 | 0.003 | 0.003 | 5.10x10^-1^ | TRUE |
| rs13057823 | A | 0.310 | 0.073 | 0.012 | 1.63x10^-9^ | 0.304 | 0.013 | 0.002 | 3.60x10^-6^ | TRUE |
| rs1321917 | C | 0.404 | 0.067 | 0.011 | 2.71x10^-9^ | 0.408 | -0.001 | 0.002 | 9.20x10^-1^ | TRUE |
| rs1913707 | A | 0.599 | 0.069 | 0.011 | 2.09x10^-9^ | 0.612 | -0.001 | 0.002 | 9.20x10^-1^ | TRUE |
| rs2078396 | T | 0.366 | -0.074 | 0.012 | 2.70x10^-10^ | 0.383 | 0.001 | 0.002 | 2.30x10^-1^ | TRUE |
| rs2268023 | A | 0.419 | 0.066 | 0.011 | 7.12x10^-9^ | 0.395 | 0.006 | 0.002 | 2.30x10^-2^ | TRUE |
| rs111844273 | A | 0.020 | 0.260 | 0.041 | 3.34x10^-10^ | 0.021 | 0.005 | 0.006 | 5.70x10^-1^ | TRUE |

*^a^refers to whether the r^2^ was greater for the SNP-exposure or the SNP-outcome relationship. If the SNP passed Steiger filtering the r^2^ was greater for the SNP-exposure association*

*Abbreviations: eBMD: estimated bone mineral density; OA: osteoarthritis; EA: effect allele; EAF: effect allele frequency; SE: standard error*

###### Supplementary Table 5: Associations between the SNPs used to instrument knee OA in 2SMR and hip OA in the GO consortium and eBMD in UK Biobank

|  |  | **Exposure= knee OA** | | | | **Outcome=eBMD** | | | |  |
| --- | --- | --- | --- | --- | --- | --- | --- | --- | --- | --- |
| **SNP** | **EA** | **EAF** | **Beta** | **SE** | **P** | **EAF** | **Beta** | **SE** | **P** | **passed steiger filtering^a^** |
| rs143384 | A | 0.586 | 0.060 | 0.009 | 5.40x10^-11^ | 0.598 | 0.003 | 0.002 | 3.30x10^-1^ | TRUE |
| rs4548913 | A | 0.623 | -0.053 | 0.009 | 6.62x10^-9^ | 0.639 | -0.025 | 0.002 | 4.50x10^-32^ | FALSE |
| rs66906321 | T | 0.172 | -0.074 | 0.012 | 5.69x10^-10^ | 0.181 | -0.015 | 0.002 | 1.10x10^-8^ | TRUE |
| rs9940278 | T | 0.443 | 0.064 | 0.009 | 6.93x10^-13^ | 0.421 | 0.019 | 0.002 | 1.60x10^-19^ | TRUE |

*^a^refers to whether the r^2^ was greater for the SNP-exposure or the SNP-outcome relationship. If the SNP passed Steiger filtering the r^2^ was greater for the SNP-exposure association*

*Abbreviations: eBMD: estimated bone mineral density; OA: osteoarthritis; EA: effect allele; EAF: effect allele frequency; SE: standard error*

###### Supplementary Table 6: Associations between the SNPs used to instrument hip OA in 2SMR and hip OA in the GWAS meta-analysis of UK Biobank and arcoGEN and BMI in the GIANT consortium

|  | | **Exposure=Hip OA** | | | | **Outcome=BMI** | | | |  |
| --- | --- | --- | --- | --- | --- | --- | --- | --- | --- | --- |
| **SNP** | **EA** | **EAF** | **Beta** | **SE** | **P** | **EAF** | **Beta** | **SE** | **p** | **passed steiger filtering^a^** |
| rs10492367 | T | 0.190 | 0.152 | 0.015 | 1.25x10^-24^ | 0.175 | -0.002 | 0.005 | 7.18x10^-1^ | TRUE |
| rs11059094 | T | 0.478 | 0.076 | 0.012 | 7.38x10^-11^ | 0.422 | 0.008 | 0.004 | 7.33x10^-2^ | TRUE |
| rs11583641 | T | 0.276 | -0.081 | 0.013 | 5.58x10^-10^ | 0.250 | 0.003 | 0.004 | 5.70x10^-1^ | TRUE |
| rs12040949 | T | 0.384 | -0.067 | 0.012 | 2.84x10^-8^ | 0.379 | 0.005 | 0.004 | 2.50x10^-1^ | TRUE |
| rs12209223 | A | 0.103 | 0.156 | 0.019 | 3.88x10^-16^ | 0.133 | -0.018 | 0.006 | 4.53x10^-3^ | TRUE |
| rs1913707 | A | 0.612 | 0.080 | 0.012 | 2.96x10^-11^ | 0.550 | -0.003 | 0.004 | 4.09x10^-1^ | TRUE |
| rs2785988 | A | 0.299 | 0.083 | 0.013 | 7.30x10^-11^ | 0.280 | 0.009 | 0.003 | 1.10x10^-2^ | TRUE |
| rs2836618 | A | 0.261 | 0.088 | 0.013 | 3.20x10^-11^ | 0.325 | 0.004 | 0.004 | 3.66x10^-1^ | TRUE |
| rs3774355 | A | 0.360 | 0.091 | 0.012 | 8.20x10^-14^ | 0.292 | 0.010 | 0.003 | 9.86x10^-4^ | TRUE |
| rs4338381 | A | 0.632 | 0.095 | 0.012 | 4.37x10^-15^ | 0.612 | 0.004 | 0.004 | 2.58x10^-1^ | TRUE |
| rs62063281 | A | 0.777 | -0.096 | 0.014 | 5.30x10^-12^ | 0.800 | -0.004 | 0.005 | 3.95x10^-1^ | TRUE |
| rs7222178 | A | 0.199 | 0.097 | 0.015 | 3.78x10^-11^ | 0.183 | 0.008 | 0.006 | 1.43x10^-1^ | TRUE |
| rs74767794 | A | 0.683 | 0.075 | 0.013 | 2.56x10^-9^ | 0.658 | 0.007 | 0.003 | 4.29x10^-2^ | TRUE |
| rs7571789 | T | 0.476 | 0.089 | 0.012 | 3.26x10^-14^ | 0.458 | 0.003 | 0.004 | 4.18x10^-1^ | TRUE |
| rs79056043 | A | 0.950 | -0.163 | 0.027 | 1.33x10^-9^ | 0.967 | 0.007 | 0.008 | 3.99x10^-1^ | TRUE |
| rs798748 | T | 0.382 | -0.072 | 0.012 | 2.50x10^-9^ | 0.308 | -0.002 | 0.004 | 5.81x10^-1^ | TRUE |
| rs80287694 | A | 0.887 | -0.109 | 0.018 | 2.66x10^-9^ | 0.883 | -0.001 | 0.006 | 8.37x10^-1^ | TRUE |

*^a^refers to whether the r^2^ was greater for the SNP-exposure or the SNP-outcome relationship. If the SNP passed Steiger filtering the r^2^ was greater for the SNP-exposure association*

*Abbreviations: BMI: body mass index; OA: osteoarthritis; EA: effect allele; EAF: effect allele frequency; SE: standard error*

###### Supplementary Table 7: Associations between the SNPs used to instrument knee OA in 2SMR and knee OA in the meta-analysis of UK Biobank and arcOGEN and BMI in the GIANT consortium

|  |  | **Exposure= knee OA** | | | | **Outcome=BMI** | | | |  |
| --- | --- | --- | --- | --- | --- | --- | --- | --- | --- | --- |
| **SNP** | **EA** | **EAF** | **Beta** | **SE** | **p** | **EAF** | **Beta** | **SE** | **p** | **passed steiger filtering^a^** |
| rs1078301 | A | 0.732 | -0.068 | 0.011 | 1.27x10^-10^ | 0.742 | -0.004 | 0.004 | 4.26x10^-1^ | TRUE |
| rs143384 | A | 0.597 | 0.094 | 0.010 | 4.77x10^-23^ | 0.600 | -0.001 | 0.003 | 8.25x10^-1^ | TRUE |
| rs4775006 | A | 0.411 | 0.058 | 0.009 | 8.40x10^-10^ | 0.422 | -0.005 | 0.005 | 2.58x10^-1^ | TRUE |
| rs8067763 | A | 0.594 | -0.057 | 0.010 | 2.39x10^-9^ | 0.600 | 0.004 | 0.004 | 2.58x10^-1^ | TRUE |
| rs8067895 | A | 0.286 | 0.062 | 0.010 | 1.89x10^-9^ | 0.292 | 0.005 | 0.004 | 2.72x10^-1^ | TRUE |
| rs9277552 | T | 0.211 | -0.064 | 0.011 | 1.97x10^-8^ | 0.275 | -0.003 | 0.004 | 5.55x10^-1^ | TRUE |

*^a^refers to whether the r^2^ was greater for the SNP-exposure or the SNP-outcome relationship. If the SNP passed Steiger filtering the r^2^ was greater for the SNP-exposure association*

*Abbreviations: BMI: body mass index; OA: osteoarthritis; EA: effect allele; EAF: effect allele frequency; SE: standard error*

###### Supplementary Table 8: descriptives of the UK Biobank population

|  | **Total population** | | | | | | | | **Multivariable MR population** | | | | | | | |
| --- | --- | --- | --- | --- | --- | --- | --- | --- | --- | --- | --- | --- | --- | --- | --- | --- |
|  | **Hip OA cases**  **N=10 525** | | **Hip OA controls**  **N=323 536** | | **Knee OA cases**  **N=18 384** | | **Knee OA controls**  **N=323 536** | | **Hip OA cases**  **N=6102** | | **Hip OA controls**  **N=184 073** | | **Knee OA cases**  **N=10 331** | | **Knee OA controls**  **N=184 073** | |
|  | **Mean** | **SD** | **Mean** | **SD** | **Mean** | **SD** | **Mean** | **SD** | **Mean** | **SD** | **Mean** | **SD** | **Mean** | **SD** | **Mean** | **SD** |
| Age, years | 61.7 | 6.0 | 56.2 | 8.1 | 60.2 | 6.9 | 56.2 | 8.1 | 61.7 | 6.0 | 56.0 | 8.0 | 60.1 | 6.9 | 56.0 | 8.0 |
| Height, cm | 167.8 | 8.9 | 168.9 | 9.2 | 168.6 | 9.4 | 168.9 | 9.2 | 167.7 | 8.8 | 168.9 | 9.2 | 168.5 | 9.4 | 168.9 | 9.2 |
| Weight, kg | 81.4 | 16.3 | 77.5 | 15.6 | 86.1 | 16.9 | 77.5 | 15.6 | 81.3 | 16.1 | 77.5 | 15.5 | 86.1 | 16.8 | 77.5 | 15.5 |
| BMI, kg/m^2^ | 28.9 | 5.0 | 27.1 | 4.6 | 30.3 | 5.4 | 27.1 | 4.6 | 28.8 | 4.9 | 27.1 | 4.6 | 30.3 | 5.4 | 27.1 | 4.6 |
| eBMD, g/cm^2^ | 0.543 | 0.148 | 0.540 | 0.134 | 0.552 | 0.151 | 0.540 | 0.134 | 0.543 | 0.148 | 0.542 | 0.134 | 0.553 | 0.151 | 0.542 | 0.134 |
|  | **N** | **%** | **N** | **%** | **N** | **%** | **N** | **%** | **N** | **%** | **N** | **%** | **N** | **%** | **N** | **%** |
| Female | 6000 | 57.0 | 174 269 | 53.9 | 9220 | 50.2 | 174 269 | 53.9 | 3460 | 56.7 | 99 127 | 53.9 | 5217 | 50.5 | 99 127 | 53.9 |

*Abbreviations: BMI: body mass index; eBMD: estimated bone mineral density; OA: osteoarthritis*

###### Supplementary Table 9: Observational relationships between eBMD, BMI and OA in the UK Biobank population

|  | | Exposure | | | |
| --- | --- | --- | --- | --- | --- |
|  |  | eBMD | BMI | Hip OA | Knee OA |
| Outcome | eBMD |  | 0.10 (0.09, 0.10) | 0.10 (0.08, 0.13) | 0.12 (0.10, 0.14) |
|  | BMI | 0.11 (0.10, 0.11) |  | 0.36 (0.34, 0.38) | 0.65 (0.63, 0.66) |
|  | Hip OA | 1.12 (1.09, 1.15) | 1.41 (1.39, 1.44) |  |  |
|  | Knee OA | 1.13 (1.11, 1.15) | 1.73 (1.71, 1.75) |  |  |

*All p<2x10^-16^*

*Effect estimates represent the SD increase in outcome per SD increase in exposure for BMD-BMI and BMI-BMD analyses, the odds ratio per SD increase in exposure for BMI-OA and BMD-OA analyses and the mean difference in SD units between those with and without hip/knee OA*

*Abbreviations: BMD: bone mineral density; BMI: body mass index*

###### Supplementary Table 10: Associations between the four genetic risk scores used as instruments and potential confounders

|  | Score | | | |
| --- | --- | --- | --- | --- |
|  | BMD | BMI | Hip OA | Knee OA |
| Age, years | -3x10^-3^ (-0.01, 3x10^-3^) | -5x10^-3^ (-0.01, 4x10^-4^) | 0.01 (-0.01, 0.02) | 5x10^-4^ (-0.02, 0.02) |
| Height, cm | 0.03 (0.02, 0.04) | 0.01 (-7x10^-5^, 0.01) | -0.01 (-0.03, 1x10^-3^) | -0.15 (-0.17, -0.13) |
| Weight, kg | 0.04 (0.03, 0.06) | 0.32 (0.31, 0.33) | -2x10^-3^ (-0.03, 0.02) | 0.33 (0.29, 0.37) |
| BMI, kg/m^2^ | 0.01 (2x10^-3^, 0.01) | *0.11 (0.11, 0.11)* | 4x10^-3^ (-4x10^-3^, 0.01) | 0.16 (0.15, 0.18) |
| eBMD, g/cm^2^ | *0.004 (0.004, 0.004)* | 2x10^-4^ (1x10^-4^, 3x10^-4^) | 5x10^-4^ (2x10^-4^, 0.001) | 2x10^-3^ (2x10^-3^, 3x10^-3^) |
| Hip OA | 1.01 (1.00, 1.01) | 1.01 (1.01, 1.02) | *1.07 (1.06, 1.08)* | 1.02 (1.00, 1.04) |
| Knee OA | 1.01 (1.00, 1.01) | 1.02 (1.01, 1.02) | 1.05 (1.02, 1.08) | *1.06 (1.05, 1.07)* |
| Physical Activity, total weekly MET-minutes | -2.82 (-7.56, 1.93) | 2.77 (-1.30, 6.84) | -0.39 (-11, 10) | -0.48 (-16, 15) |
| HRT use, ever | 1.00 (1.00, 1.00) | 1.00 (1.00, 1.00) | 1.00 (1.00, 1.01) | 1.00 (0.99, 1.01) |

*Associations between the scores and the exposure they were used to instrument are also included for comparison of magnitude of effect. Estimates for continuous covariates represent the per-allele unit increase (e.g. a value of 0.03 for height represents a per-allele increase in height of 0.03cm). Estimates for binary variables represent the per-allele odds ratio.*

###### Supplementary Table 11: results of two-sample MR analyses

|  | |  |  |  | **IVW** | | **MR Egger** | | | | **Weighted median** | |
| --- | --- | --- | --- | --- | --- | --- | --- | --- | --- | --- | --- | --- |
| **Exposure** | | **Steiger filtered SNPs** | **N SNPs^a^** | **Outcome** | **estimate (95% CI)** | ***p* value** | **estimate (95% CI)** | ***p* value** | **intercept** | ***p* value** | **estimate (95% CI)** | ***p* value** |
| eBMD | | 7 | 346 | Hip OA | 1.09 (1.03, 1.16) | 0.002 | 1.04 (0.95, 1.14) | 0.416 | 1.00 (1.00, 1.01) | 0.178 | 1.05 (0.97, 1.15) | 0.236 |
|  | *Excluding BMI Steiger filtered SNPs* | 2 | 344 | Hip OA | 1.09 (1.03, 1.16) | 0.002 | 1.04 (0.94, 1.14) | 0.450 | 1.00 (1.00, 1.01) | 0.156 | 1.05 (0.96, 1.15) | 0.264 |
|  | *Restricted to SNPs associated with FN-BMD at genome-wide significance* | 0 | 10 | Hip OA | 1.40 (1.12, 1.74) | 0.002 | 0.89 (0.37, 2.10) | 0.789 | 1.03 (0.98, 1.07) | 0.313 | 1.22 (0.98, 1.52) | 0.068 |
| eBMD | | 4 | 349 | Knee OA | 1.04 (1.00, 1.09) | 0.068 | 1.07 (0.99, 1.16) | 0.086 | 1.00 (1.00, 1.00) | 0.554 | 1.02 (0.94, 1.10) | 0.691 |
|  | *Excluding BMI Steiger filtered SNPs* | 2 | 347 | Knee OA | 1.04 (1.00, 1.09) | 0.067 | 1.07 (0.99, 1.16) | 0.090 | 1.00 (1.00, 1.00) | 0.479 | 1.02 (0.94, 1.09) | 0.687 |
|  | *Restricted to SNPs associated with FN-BMD at genome-wide significance* | 0 | 10 | Knee OA | 1.21 (1.01, 1.44) | 0.034 | 1.15 (0.54, 2.41) | 0.730 | 1.00 (0.96, 1.04) | 0.886 | 1.26 (1.07, 1.49) | 0.006 |
| eBMD | | 2 | 267 | BMI | 0.01 (-0.01, 0.03) | 0.472 | 3x10^-3^ (-0.03, 0.04) | 0.877 | 2x10^-4^ (-9x10^-4^, 1x10^-3^) | 0.737 | 0.02 (-0.01, 0.05) | 0.209 |
|  | *Excluding hip and knee OA Steiger filtered SNPs* | 9 | 258 | BMI | 3x10^-3^ (-0.02, 0.02) | 0.771 | 5x10^-3^ (-0.03, 0.04) | 0.802 | -6x10^-5^ (-1x10^-3^, 1x10^-3^) | 0.919 | 0.02 (-0.01, 0.05) | 0.231 |
| BMI | | 4 | 58 | Hip OA | 1.56 (1.31, 1.87) | 8x10^-7^ | 2.01 (1.31, 3.08) | 0.002 | 0.99 (0.98, 1.00) | 0.208 | 1.83 (1.50, 2.21) | 1x10^-9^ |
|  | *Excluding eBMD Steiger filtered SNPs* | 8 | 50 | Hip OA | 1.46 (1.26, 1.71) | 1x10^-6^ | 2.01 (1.41, 2.84) | 3x10^-4^ | 0.99 (0.98, 1.00) | 0.056 | 1.74 (1.44, 2.11) | 1x10^-8^ |
| BMI | | 1 | 61 | Knee OA | 1.69 (1.48, 1.93) | 1x10^-14^ | 1.41 (1.02, 1.95) | 0.040 | 1.01 (1.00, 1.02) | 0.228 | 1.67 (1.44, 1.95) | 2x10^-11^ |
|  | *Excluding eBMD Steiger filtered SNPs* | 10 | 51 | Knee OA | 1.61 (1.42, 1.84) | 1x10^-12^ | 1.40 (1.03, 1.90) | 0.037 | 1.00 (1.00, 1.01) | 0.317 | 1.67 (1.43, 1.96) | 3x10^-10^ |
| BMI | | 11 | 51 | eBMD | 0.13 (0.09, 0.17) | 7x10^-10^ | 0.23 (0.14, 0.33) | 7x10^-6^ | -0.01 (-0.01, -4x10^-4^) | 0.035 | 0.17 (0.14, 0.21) | 8x10^-19^ |
|  | *Excluding hip and knee OA Steiger filtered SNPs* | 3 | 48 | eBMD | 0.13 (0.09, 0.17) | 1x10^-9^ | 0.24 (0.15, 0.33) | 9x10^-6^ | -4x10^-3^ (-0.01, -1x10^-3^) | 0.016 | 0.18 (0.14, 0.22) | 2x10^-19^ |
| Hip OA | | 0 | 9 | eBMD | 0.02 (-0.01, 0.05) | 0.117 | -1x10^-3^ (-0.09, 0.09) | 0.976 | 2x10^-3^ (-0.01, 0.01) | 0.622 | 0.01 (-0.01, 0.03) | 0.396 |
| Hip OA | | 0 | 17 | BMI | 0.02 (-5x10^-3^, 0.04) | 0.120 | -0.07 (-0.16, 0.01) | 0.110 | 0.01 (9x10^-4^, 0.02) | 0.046 | 0.02 (7x10^-4^, 0.05) | 0.044 |
|  | *excluding*  *rs12209223^b^* | 0 | 16 | BMI | 0.03 (0.01, 0.04) | 0.009 | -0.03 (-0.11, 0.06) | 0.548 | 4.9  5x10^-3^ (-3x10^-3^, 0.01) | 0.242 | 0.03 (4x10^-3^, 0.05) | 0.023 |
| Knee OA | | 1 | 3 | eBMD | 0.13 (0.03, 0.23) | 0.010 | 0.53 (-0.99, 2.06) | 0.617 | -0.03 (-0.12, 0.07) | 0.696 | 0.10 (0.06, 0.14) | 5x10^-6^ |
| Knee OA | | 0 | 6 | BMI | 1x10^-4^ (-0.03, 0.03) | 0.995 | 0.02 (-0.15, 0.19) | 0.842 | -1x10^-3^ (-0.01, 0.01) | 0.839 | 5x10^-3^ (-0.03, 0.04) | 0.821 |

*^a^ Number of SNPs after Steiger filtering*

*^b^ The rs12209223 OA-risk increasing allele was identified in a GWAS of height, with the same allele associated with increased height*

*Effect sizes for binary exposures represents the SD increase per doubling in OA risk. Effect sizes for BMD and BMI analyses represent the SD increase in outcome per SD increase in exposure.*

*Abbreviations: IVW: inverse variance weighted; SNPs: single nucleotide polymorphisms; eBMD: estimated bone mineral density; FN-BMD: femoral neck bone mineral density; BMI: body mass index; OA: osteoarthritis*

*Supplementary Figure 1: plot of two-sample MR results for the relationship between estimated bone mineral density (eBMD) and hip osteoarthritis (OA)*

###### Supplementary Table 12: Results of latent causal variable modelling for the genetic correlation between femoral neck/ lumbar spine BMD and hip/knee OA, as well as the estimate of the genetic causality proportion

| **Trait 1** | **Trait 2** | **Rho (SE)** | **GCP (SE)** | ***P* value** |
| --- | --- | --- | --- | --- |
| Femoral neck BMD | Hospital-diagnosed hip OA | 0.16 (0.02) | 0.56 (0.07) | 3x10^-20^ |
|  | Hospital-diagnosed knee OA | 0.19 (0.07) | 0.64 (0.21) | 1x10^-7^ |
| Lumbar spine BMD | Hospital-diagnosed hip OA | 0.23 (0.08) | 0.57 (0.21) | 0.002 |
|  | Hospital-diagnosed knee OA | 0.20 (0.07) | 0.59 (0.25) | 0.003 |

*Rho represents the genetic correlation between the two traits, estimated by LD score regression. The GCP is an estimate of the genetic component of trait 1 which is causal for trait 2. A value closer to 1 represents stronger causality of trait 1 on trait 2. A negative value represents the proportion of the genetic component for trait 2 that is causal for trait 1 (15)*

#### Supplementary Figures

Supplementary Figure 1: plot of two-sample MR results for the relationship between estimated bone mineral density (eBMD) and hip osteoarthritis (OA)
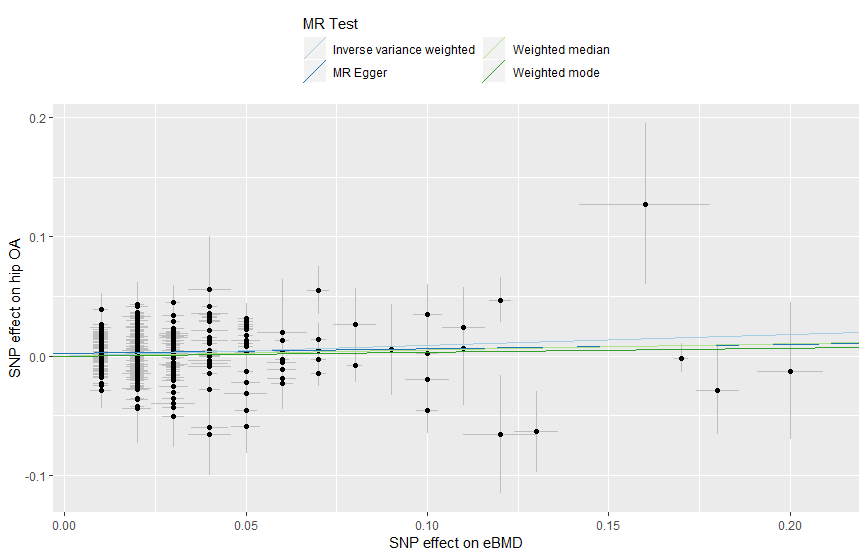

Supplementary Figure 2: plot of two-sample MR results for the relationship between estimated bone mineral density (eBMD) and hip osteoarthritis (OA)
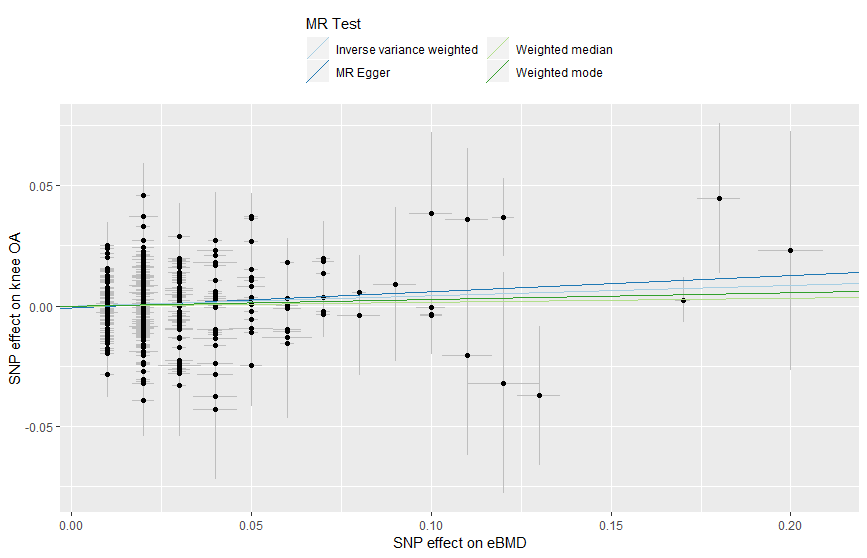

Supplementary Figure 3: plot of two-sample MR results for the relationship between estimated hip osteoarthritis (OA) and bone mineral density (eBMD)

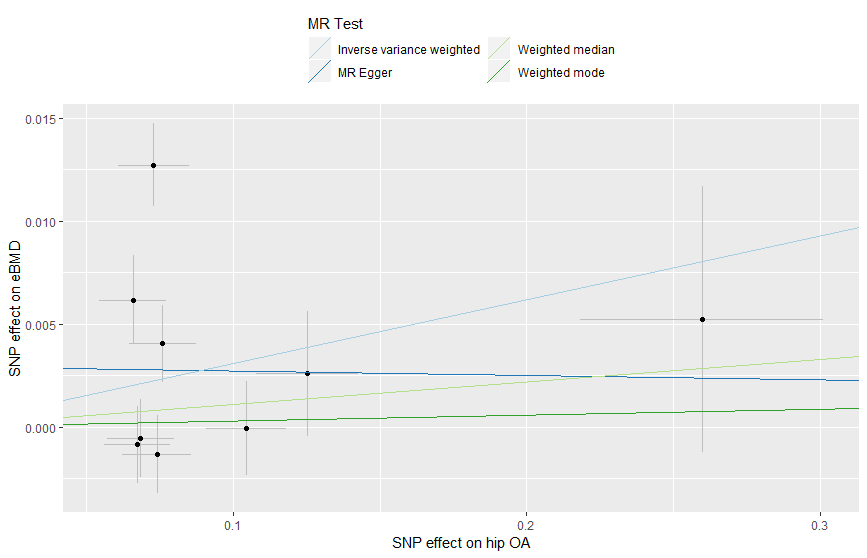

###### Supplementary Figure 4: plot of two-sample MR results for the relationship between knee osteoarthritis (OA) and estimated bone mineral density (eBMD)

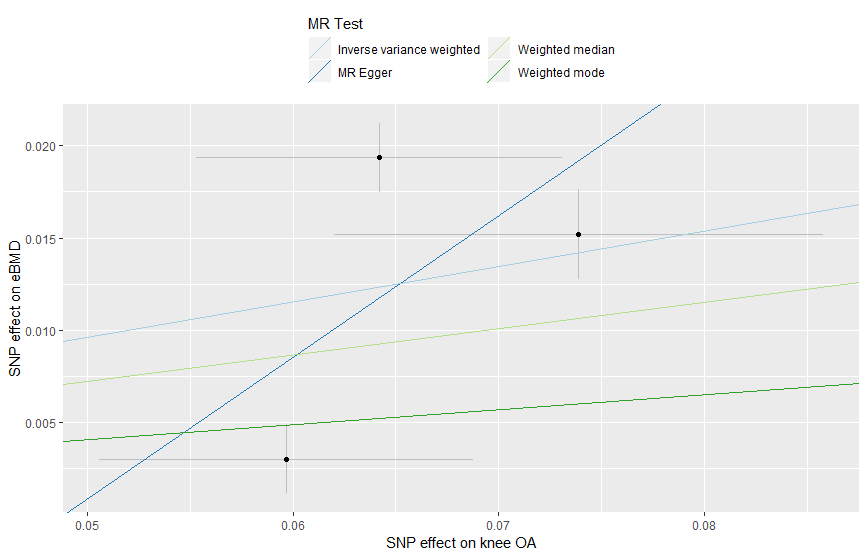

###### Supplementary Figure 5: plot of two-sample MR results for the relationship between body mass index (BMI) and hip osteoarthritis (OA)

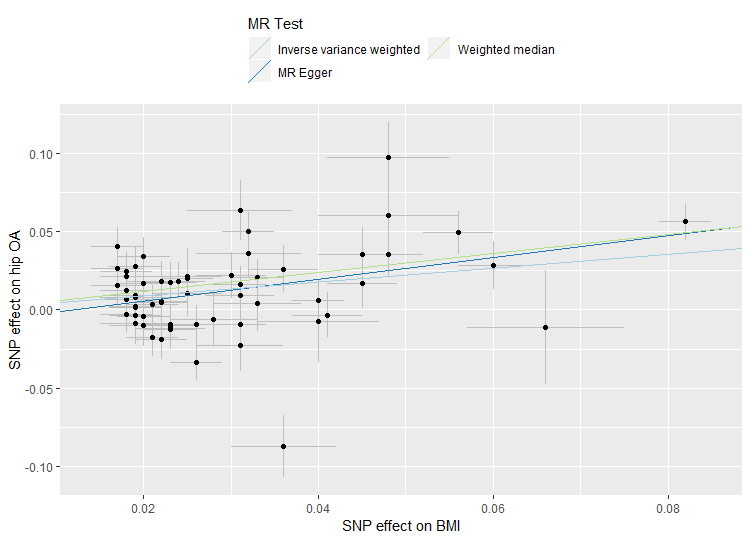

###### Supplementary Figure 6: plot of two-sample MR results for the relationship between body mass index (BMI) and knee osteoarthritis (OA)

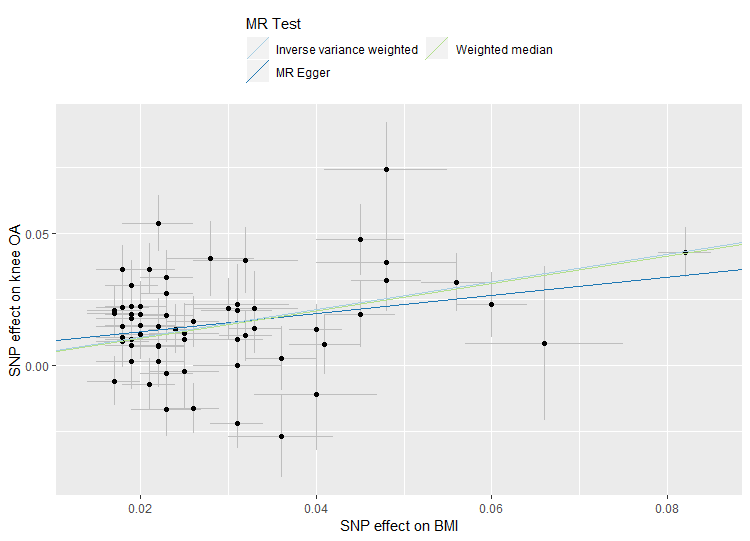

###### Supplementary Figure 7: plot of two-sample MR results for the relationship between hip osteoarthritis (OA) and body mass index (BMI)

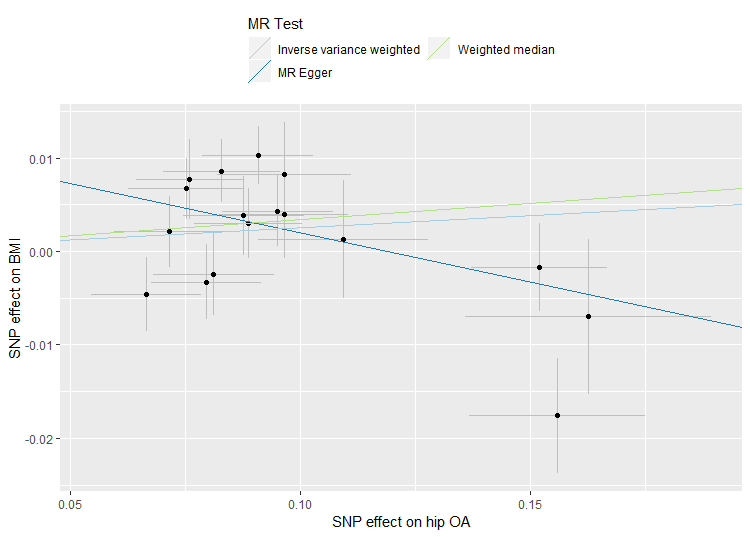

###### Supplementary Figure 8: plot of two-sample MR results for the relationship between knee osteoarthritis (OA) and body mass index (BMI)

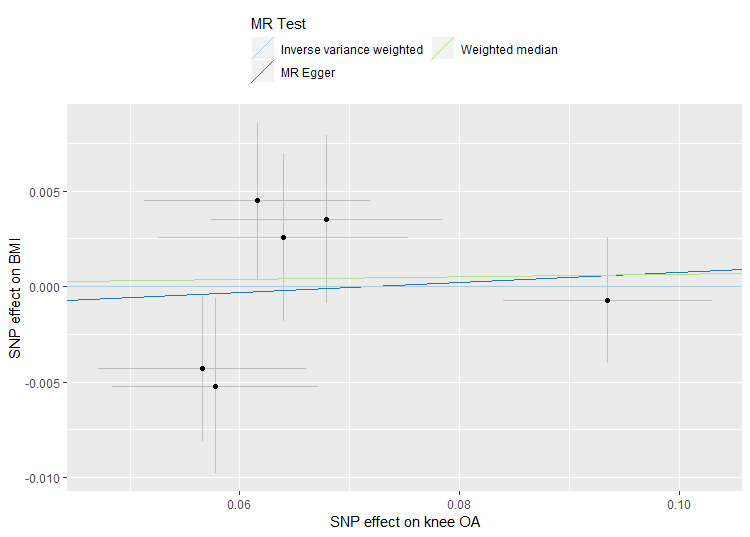

###### Supplementary Figure 9: plot of two-sample MR results for the relationship between estimated bone mineral density (eBMD) and body mass index (BMI)

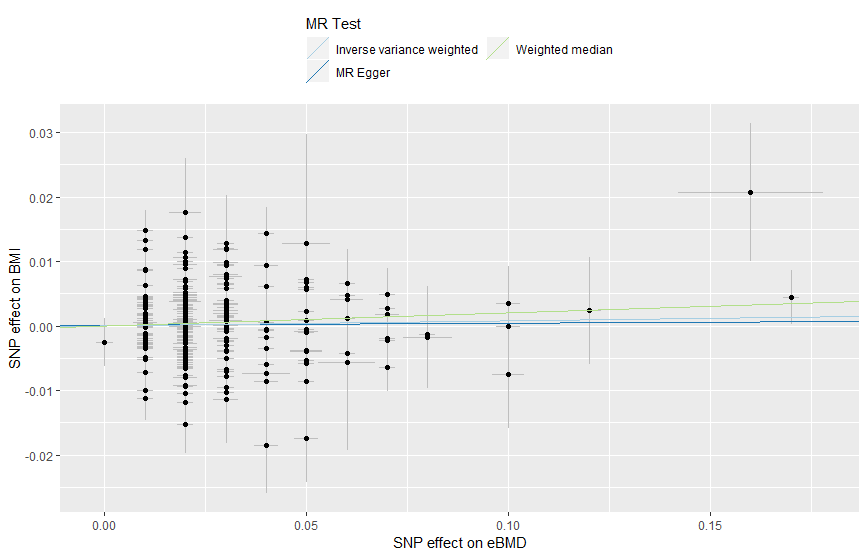

###### Supplementary Figure 10: plot of two-sample MR results for the relationship between body mass index (BMI) and estimated bone mineral density (eBMD)

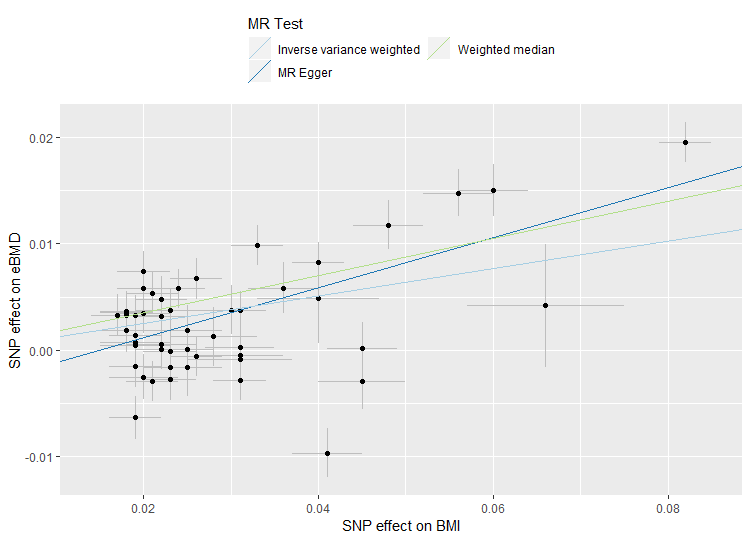

###### Supplementary Figure 11: quantile-quantile (qq) plots of p values from the weight-adjusted GWAS of hospital-diagnosed hip OA (a) and hospital-diagnosed knee OA (b) in UK Biobank

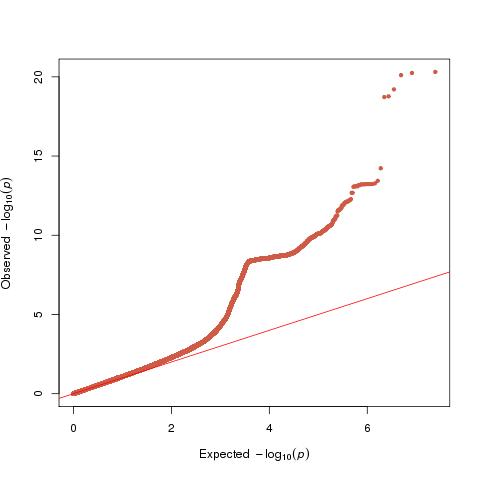

**(a)**

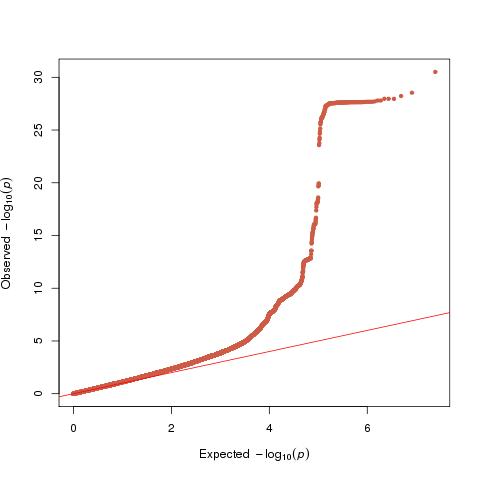

**(b)**

*λ_hip_=1.05, λ_knee_=1.09*
